# The Impact of Delayed School Entry on Developmental Outcomes in Very Preterm Infants

**DOI:** 10.1101/2025.10.29.25339058

**Authors:** Anna Haefke, Antonia Schedewi, Carla Mehring, Wolfgang Goepel, Stephanie Weibel, Juliane Spiegler

## Abstract

**Objective:** To investigate whether delayed school entry (DSE) benefits outcomes related to everyday functioning and participation in very preterm-born children.

**Methods:** We conducted a systematic review according to Cochrane standard. Additionally, we analyzed data of 1620 children born before 32 weeks of gestation at five and ten years from a prospective cohort study. Children with DSE were compared to those with age-appropriate school entry (ASE) regarding everyday functioning and participation, using standardized parent and self-report questionnaires.

**Results:** Our systematic review identified one study reporting no benefit of DSE on academic skills (very low certainty of evidence). In the prospective cohort, DSE was associated with less favorable outcomes: participation (ASE 95.8±8.5; DSE 90.3±14.0; p<0.001), attention (ASE 3.6±1.2; DSE 4.1±1.3; p<0.001), internalizing problems (ASE 4.5±3.3; DSE 5.7±4.0; p=0.01), executive functioning (ASE 52±11; DSE 58±13; p<0.001) but not in quality of life (ASE 54.0±6.2; DSE 52.9±6.4; p=0.17). DSE children had higher special educational needs (34% vs. 9.8%; p<0.001). However, after adjustment for predefined confounders, no significant differences remained at ten years.

ASE based on chronological age was associated with delayed transition to secondary school in contrast to ASE based on corrected age.

**Conclusion:** After adjustment, DSE had no effect on outcomes relating to everyday functioning and participation at ten years or the timely transition to secondary school. Age corrected for preterm birth should be used to determine school entry age. Further research is needed on the long-term academic implications of age at school entry.

## Introduction

Preterm birth, (delivery < 37 weeks of gestation), remains a significant challenge for healthcare systems. In Germany, 674,965 children were born in 2023, including 53,137 preterm births. Of these 9,630 were very preterm children (VPT, gestational age < 32 weeks).^1^ VPT face higher risk of medical complications, neurological impairments, cognitive deficits, attention problems, emotional and social difficulties.^2^

Such challenges frequently persist into school years, with a higher incidence of learning difficulties, limited attention regulation and greater need for educational support.^3,4^ Compulsory education in Germany begins at the age of six years, with cut-off date for the age ranging from June 30 to September 30 depending on the federal state. Children deemed immature during school entry assessments, may be deferred to the following academic year.

Larsen et al. examined the relationship between delayed school entry (DSE) and academic achievement of term-born infants. Minor advantage in reading and numeracy performance for DSE were found in grades three, five and seven, but equalized by grade nine.^5^ After adjusting for confounders, including inattention and hyperactivity, DSE showed no consistent benefits compared with age-appropriate school entry (ASE) at any grade level. Although conducted in Australia on term-born children, the study provides relevant insights.

Given the substantial evidence gap, the present study was conducted using the F-words- model to analyze meaningful outcomes for VPT. Based on the World Health Organizations International Classification of Functioning, Disability and Health, this model highlights six key dimensions of child development: *Function, Friendship (social participation), Family, Fun, Fitness (Physical health), Future.*^6^

The objective of this investigation is to examine whether DSE has any benefits on outcomes across key domains of the F-words framework for VPT. The second objective was to test whether using chronological or age corrected for prematurity changes outcomes for VPT.

## Methods

### Systematic review

As part of the ongoing update and upgrade of the German evidence-based guideline for the follow-up of preterm infants, the question of whether DSE may be beneficial for preterm children has been raised. In this context, our guideline working group conducted a systematic literature review. Details on methods and search strategy are provided in Appendices 1 and 2.

### Cohort study: Design and Setting

We analyzed a prospective cohort study of 1620 preterm infants (gestational age <32 weeks and birthweight <1,500g) across hospitals throughout Germany that participate in the German Neonatal Network (GNN).

The GNN is a prospective, multicenter cohort study that has been enrolling very low birth weight infants (<1500g) since 1^st^ January 2009. Recruiting occurred immediately after birth, and perinatal data were collected before discharge. At the ages of five and ten years, additional data on the child’s health and development were gathered through parent- and self-reported questionnaires. At the age of five years, a comprehensive developmental assessment was conducted.

For this analysis the sample was restricted exclusively to children born between 2009 and 2016 as follow-up data from the five- and ten-years questionnaires were available for this cohort. We excluded participants with an intelligence quotient (IQ) below 70 at the age of five years, as they most likely enter a specialized school system in Germany.

Potential confounders were selected based on the literature ^2,7–9^ with neonatal confounders (gestational age, birth weight, male sex, Bronchopulmonary Dysplasia (BPD), intraventricular hemorrhage (IVH)), mother’s high school diploma as proxy for socioeconomic status) and with confounders measured before school entry (diagnosis of cerebral palsy, club sport participation, IQ (Wechsler Preschool and Primary Scale III), Strengths and Difficulties Questionnaire (SDQ), Movement Assessment Battery for Children-II) .

### Testing Methods and Tools

The following data were inquired through standardized parental or self-reported questionnaires during the ten-year follow-up examination and assigned to the corresponding dimensions of the F-words: *Function, Friendship, Family, Fun, Fitness and Future*.

#### Function

Special educational needs (SEN) were defined as either an individualized education program or visiting specialized schools.

Executive functions were measured by parental report using the Global Executive Composite of the Behavior Rating Inventory of Executive Function (BRIEF)^10^ as an overall score. T-scores above 65 were classified as clinically relevant.

The parental questionnaire for the Assessment of Cognitive Problems in Children and Adolescents (KOPKIJ)^11^ was utilized to evaluate cognitive functioning. The KOPKIJ gathers performance across multiple cognitive domains (language, memory, visual-spatial processing, behavioral regulation, attention, school achievement). Higher scores indicate more problems with scores above the 85^th^ percentile interpreted as clinically relevant.

Attention was measured using the attention-related questions of the SDQ (self-report of Item 15 “I am easily distracted. I find it difficult to concentrate” and Item 25 “I finish the work I’m doing. My attention is good”).^12–14^ To identify potential internalizing problems, the emotional and peer problems subscales of the SDQ were combined and used.^15^

*Friendship:* “Peer”- and “Bullying”-sub score of the self-report of the KIDSCREEN-52^16^ was used.

*Family:* “Parents”-sub score of the self-report of the KIDSCREEN-52^16^ was used. *Fun:* “Autonomy”-sub score of the parental report of the KIDSCREEN-52^16^ was used. *Fitness:* Participation in sports was defined as being a member of a sports club.

*Future:* The data set included all VPT who had been enrolled in school for a minimum of four years at the time of follow-up participation. The ASE-group was subdivided into early school entry (ESE, < age 6 years at school entry) and ASE (age 6-7 years at school entry). The prevailing school structure was classified into three categories (primary, secondary and other) and included in a comparative analysis.

#### Overall

Participation was assessed using the parental report of the Child and Adolescent Scale of Participation (CASP)^17,18^, a validated instrument covering four domains: home, school, community, and home/community-based activities. Higher scores indicate better participation. Average scores expressed as Percent of Maximum Possible (POMP) below 97.50% reflect at least “mild” participation restriction.

The health-related quality of life was assessed using the self-report of the KIDSCREEN- 52^16^, a validated questionnaire covering physical well-being, psychological well-being, social support and peers, parental relationships, autonomy and school environment. Item scores were transformed into T-scores (mean value 50, standard deviation (SD) 10), with scores below 40 (1 SD) indicating impaired quality of life. An overall score was used as well as domain-specific sub scores corresponding to the F-words framework (see above).

### School entry

Both the corrected and the chronological ages of preterm children at school entry were calculated using the school entry year and a cut-off date of June 30. Corrected age was used for all analysis as there is no rationale why brain maturation of VPT should be enhanced.^19^

DSE was defined as school entry at the age of seven years or older. ASE was defined as a school start before the 7^th^ birthday.

### Statistics

Statistical analysis was performed with IBM SPSS Vers. 29.^20^ Baseline characteristics and outcomes were described using descriptive statistics and compared using the Chi-square- and the Mann–Whitney-U-test. Subsequently, logistic and linear regression analyses were performed including relevant confounders. To correct for multiple testing, the Bonferroni correction was applied within each table. ^21^ Statistical significance was defined as p<0.0042 in Table 2, p<0.0033 in Tables 3 and 4 and p<0.017 in Figure 1, based on the number of comparisons.

**Figure 1.**
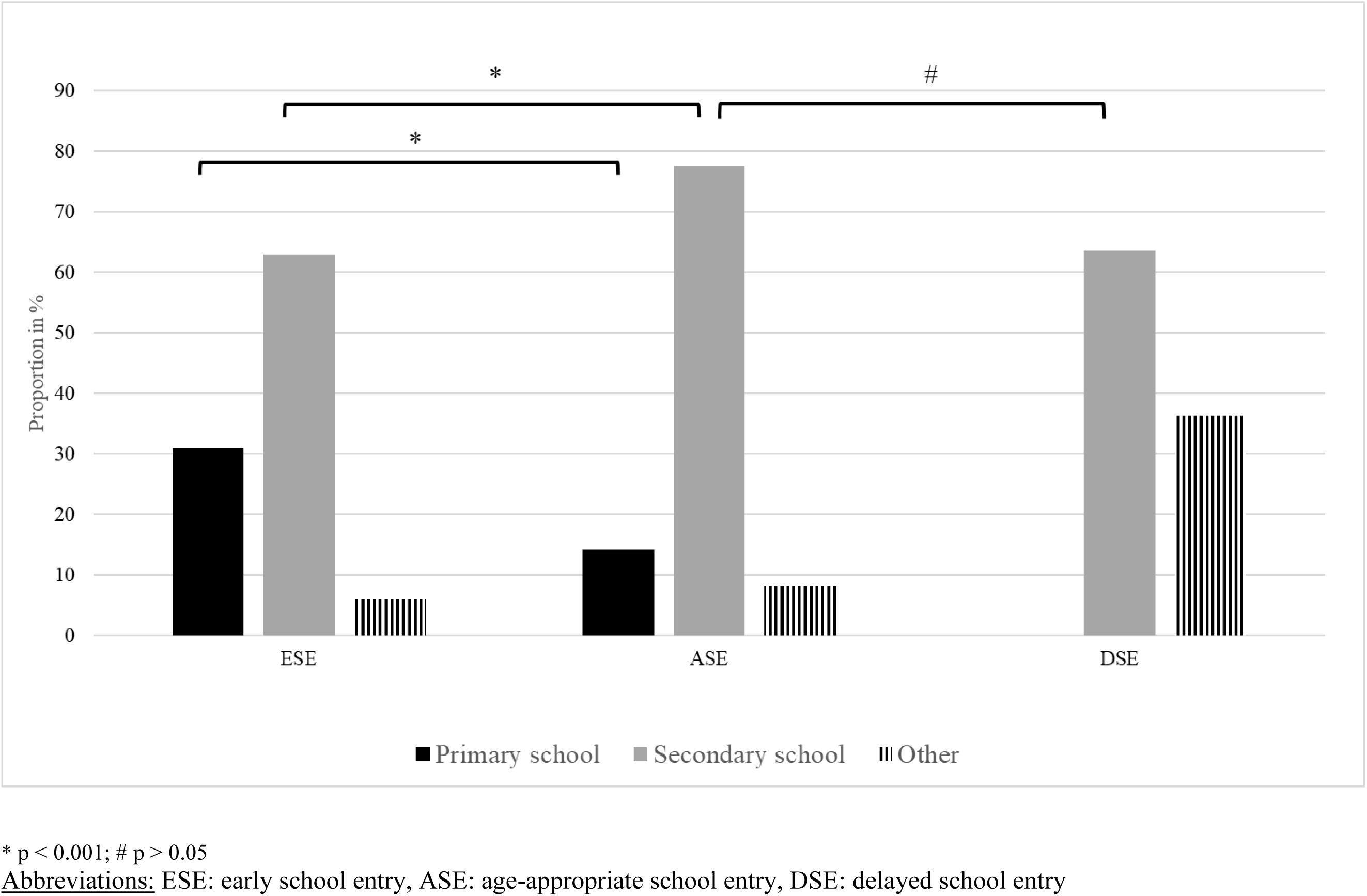
*Future:* School type after four years in school

**Table 1.** Certainty of evidence

| **Outcome**<br>*Age of assessment* | **Assessment tool** | **Number of participants**<br>*trials* | **Absolute Effect**<br>*Median* |  | **Certainty of Evidence** |
| --- | --- | --- | --- | --- | --- |
|  |  |  | Comparator | Intervention |  |
| **School performance-Reading**<br>*8 years* | Zürich Reading Test and pseudoword reading test | 340<br>(1) | 99.9 | 85.6 | Very low due to imprecision |
| **School performance-Writing**<br>*8 years* | Diagnostic spelling test | 340<br>(1) | 100.5 | 85.8 | Very low due to imprecision |
| **School performance-Mathematics**<br>*8 years* | Standardized mathematical test | 340<br>(1) | 97.5 | 88.3 | Very low due to imprecision |
| **Participation** |  |  |  |  | No evidence identified |
| **Quality of life - multi-dimensional** |  |  |  |  | No evidence identified |
| **Quality of life - Bullying** |  |  |  |  | No evidence identified |
| **Behavior - Externalizing problems: Attention**<br>*8 years* | task orientation sub-scale of the Tester's Rating of Child Behaviour | 340<br>(1) | 100.1 | 83.8 | Very low due to imprecision |
| **Behavior - Internalizing problems** |  |  |  |  | No evidence identified |
| **Behavior - Adaptive skills** |  |  |  |  | No evidence identified |
| **Social life** |  |  |  |  | No evidence identified |
| **Atypicality** |  |  |  |  | No evidence identified |
| **School problems** |  |  |  |  | No evidence identified |
| Outcomes reflect the predefined domains of interest for the systematic review. For several domains, no eligible studies were identified, indicating a substantial evidence gap. |  |  |  |  |  |

**Table 2.** Baseline characteristics

|  |  | **ASE** | **DSE** | **p-Value** |
| --- | --- | --- | --- | --- |
|  |  | 1535 (94.8%) | 85 (5.2) |  |
| **Gestational age** (weeks) | Mean (sd) | 28.0 (2.0) | 26.9 (2.1) | <0.001<sup>b</sup> |
| **Birth weight** (grams) |  | 1025 (277) | 861 (268) | <0.001<sup>b</sup> |
| **Male** | n (%) | 758 (49.4) | 53 (62) | 0.02<sup>a</sup> |
| **BPD** |  | 233 (15.2) | 29 (34) | <0.001<sup>a</sup> |
| **IVH** |  | 253 (16.5) | 24 (28) | 0.005<sup>a</sup> |
| grade I |  | 117 (7.6) | 10 (12) | 0.06<sup>a</sup> |
| grade II |  | 71 (4.6) | 8 (9) |  |
| grade III |  | 32 (2.1) | 2 (2) |  |
| grade IV |  | 33 (2.1) | 4 (5) |  |
| **Cerebral palsy** |  | 37 (2.5) | 1 (1) | 0.47<sup>a</sup> |
| **Mother high school** |  | 813 (55.6) | 36 (44) | 0.04<sup>a</sup> |
| **IQ** at age 5 | Mean (sd) | 102 (12) | 92 (11) | <0.001<sup>b</sup> |
| **SDQ** at age 5 |  | 8.6 (5.2) | 11.3 (6.1) | <0.001<sup>b</sup> |
| **MABC** at age 5 |  | 9.3 (3.0) | 7.2 (3.1) | <0.001<sup>b</sup> |
| **Club sport** at age 5 | n (%) | 876 (57.1) | 33 (39) | <0.001<sup>a</sup> |
| <sup>a</sup> tested by Chi-square test.<br><sup>b</sup> tested by Mann-Whitney U test.<br><u>Abbreviations</u><br>ASE: age-appropriate school entry, DSE: delayed school entry, BPD: Bronchopulmonary Dysplasia, IQ: Intelligence Quotient, IVH: Intraventricular Hemorrhage, MABC: Movement Assessment Battery for Children- II, SDQ: Strengths and Difficulties Questionnaire, mean: mean value, sd: standard deviation, n number |  |  |  |  |

**Table 3.**
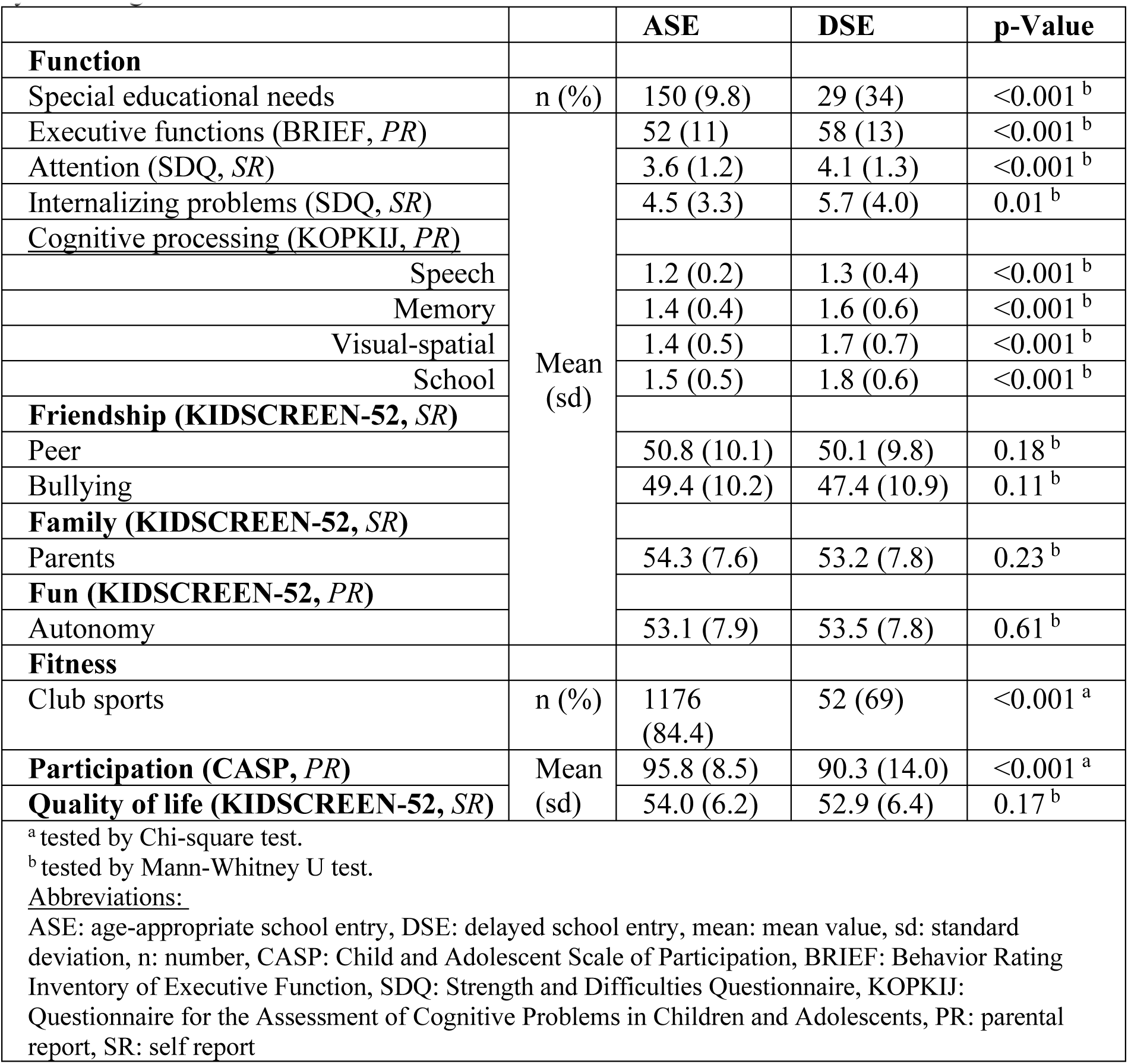
Outcomes of appropriate versus delayed school entry with regard to F-words at 10 years of age

**Table 4.** Effects of Delayed School entry (DSE)

|  | B | 95%-CI |  | p-Value |
| --- | --- | --- | --- | --- |
| Function |  |  |  |  |
| Executive functions (BRIEF, PR) | 0.573 | -1.980 | 3.126 | 0.66 |
| Attention (SDQ, SR) | 0.077 | -0.251 | 0.406 | 0.64 |
| Internalizing problems (SDQ, SR) | 0.156 | -0.773 | 1.084 | 0.74 |
| Cognitive processing (KOPKIJ, PR) |  |  |  |  |
| Speech | 0.072 | 0.008 | 0.136 | 0.03 |
| Memory | 0.048 | -0.059 | 0.156 | 0.38 |
| visual-spatial | 0.118 | -0.007 | 0.244 | 0.06 |
| School | 0.120 | -0.021 | 0.261 | 0.10 |
| Friendship (KIDSCREEN-52, SR) |  |  |  |  |
| Peers | 0.213 | -2.698 | 3.125 | 0.89 |
| Bullying | 0.451 | -2.629 | 3.530 | 0.77 |
| Family (KIDSCREEN-52, SR) |  |  |  |  |
| Parents | 0.002 | -2.309 | 2.313 | 0.99 |
| Fun (KIDSCREEN-52, PR) |  |  |  |  |
| Autonomy | 2.764 | 0.459 | 5.069 | 0.02 |
| Participation (CASP, PR) | -1.037 | -2.997 | 0.923 | 0.30 |
| Quality of life (KIDSCREEN-52, SR) | 0.716 | -1.088 | 2.519 | 0.44 |
|  | OR | 95%-CI |  | p-Value |
| Function |  |  |  |  |
| Special educational needs | 1.197 | 0.544 | 2.637 | 0.66 |
| Fitness |  |  |  |  |
| Club Sports | 0.707 | 0.319 | 1.569 | 0.40 |
| Abbreviations:<br>DSE: delayed school entry, CASP: Child and Adolescent Scale of Participation, BRIEF: Behavior Rating Inventory of Executive Function, SDQ: Strength and Difficulties Questionnaire, KOPKIJ: Questionnaire for the Assessment of Cognitive Problems in Children and Adolescents, B: regression coefficient, CI: confidence interval, OR: Odds ratio, PR: parental report, SR: self report<br>Linear and logistic regression, adjusted for: gestational age, birth weight, gender, bronchopulmonary dysplasia, intraventricular hemorrhage, mother high school, cerebral palsy, at age of 5: IQ, Movement Assessment Battery for Children, Strength and difficulties Questionnaire, club sports |  |  |  |  |

### Ethics

Approval by the local institutional review board for research in human subjects of the University of Lübeck (#08-022, #14-220, #20-334) and by the local ethic committees of all participating centers has been given.

## Results

### Systematic review

A total of 2,080 studies were screened. Some studies were thematically appropriate, but excluded due to different school structures and school entry policies, rendering them incomparable to the German context (Appendix 3, Table S3). Only one study met all inclusion criteria.^22^ This study examined academic and behavioral outcomes of preterm children born in 1985-86 in a subgroup analysis based on school entry timing in Bavaria, Germany. At age eight, children with DSE performed worse academically than those with ASE, despite having eight months less schooling at testing; differences remained after adjusting for schooling duration. Teacher ratings after the first school year showed no group differences in academic performance or attention. The certainty of evidence for all school performance domains and attention was rated as very low due to imprecision (Table 1). Consequently, the evidence regarding the effect of DSE on the outcomes remained unclear. Further details about the results, as well as included and excluded studies, are provided in Appendix 3.

### Cohort study: Baseline

Between 2009 and 2016, 13,848 VPT were included in the database. Due to financial constraints a random sub-sample of 3,403 was followed-up at age five years and eligible for follow-up at age ten years. The sampling procedure for the follow-up at five to six years has been described before explaining why only a subgroup could be followed.^23^ Of these, 1,836 (53.9%) completed both follow-up assessments. Infants with an IQ below 70 were excluded (n=216), leaving 1,620 (47.6%) VPT for analyzing. Compared to the initial GNN cohort, this sample had a significantly lower gestational age (27.8 vs. 28.0 weeks), lower birth weight (993 vs. 1015 grams) and higher incidence of BPD (20.1% vs. 18.9%). However, differences were of minor clinical importance amounting to two days or 22 grams. Children of mothers with non-German heritage were less likely to attend follow-up (12.3% vs. 27.2%; Appendix 4, Table S6).

The results presented in the following sections pertain to DSE based on age corrected for prematurity, the analysis of DSE based on the chronological age yielded only minor differences with regard to the baseline and F-words (Appendix 4, Table S7-9).

Baseline characteristics are displayed in Table 2. The DSE group included 85 (5.2%) children, born at significantly lower gestational age and birth weight. 62% children were male.

BPD, IVH and severe IVH (≥ grade III) were more prevalent in the DSE group, whereas cerebral palsy rates did not differ. Significantly, fewer mothers had a high school diploma (Table 2).

At the age of five years, the DSE group demonstrated poorer cognitive and motor development, higher SDQ-scores, suggesting a higher rate of behavior problems, and lower participation rates in club sports at the age of five years (Table 2).

### F-words

#### Function

One third of VPT with DSE received SEN (Table 3). Scores of BRIEF, the attention scale and the SDQ internalizing problems scales were significantly higher indicating more problems (Table 3). Additionally, the DSE group demonstrated significantly poorer average performances in the domains of cognitive processing (KOPKIJ) (Table 3).

#### Friendship, Family, Fun

No significant differences were observed in the overall KIDSCREEN-52 T-score, nor in the KIDSCREEN-52 dimensions corresponding to the F-words framework. (Table 3).

#### Fitness

VPT with DSE were less likely to participate in club sports than the ASE group (Table 3).

DSE was associated with significantly lower participation rates in the CASP-POMP.

Accordingly, participation in the group with ASE is “mildly” and with DSE “somewhat limited”.

However, after adjustment for confounding factors, no significant effect on the outcome at ten years could be determined (Table 4, Appendix 4 and 5).

#### Future: Timely transition to secondary school

At the time of the ten-years-questionnaires, 678 children had been at school for a minimum of four years and were typically supposed to enter secondary school. However, 1.6% of the children entered secondary school delayed.

None of the VPT with DSE were in primary school for more than the regular four years, and 14% of the VPT with ASE entered secondary school delayed (Figure 1). In contrast every third VPT with ESE stayed in primary school longer than expected.

After adjustment for confounders, DSE did not significantly affect the likelihood of the timely transition secondary school. By comparison, ASE was associated with a 2.8-fold higher chance of timely transition relative to ESE (Figure 1, Appendix 6 and 7).

## Discussion

The question of whether VPT should enter school later has been considered highly relevant by experts of the German guideline committee. However, the systematic literature review identified only one eligible study, which found no benefit of DSE on academic performance or attention. The certainty of evidence is accordingly very low. Potential effects on broader developmental domains described by the F-words framework were not reported. Our cohort study addresses this gap.

In a large German cohort, DSE did not affect the participation or quality of life of VPT considering the F-words. Very preterm birth does not enhance brain maturation; therefore, the corrected age should be used for school entry decisions. In most cases VPT have a corrected age of at least two months less than the chronological age, resulting in the probability needing more than four years for completion of primary school if the chronological age is used. This goes along with a necessary change of the year group for the VPT in most school systems in Germany.

Approximately 5% of the VPT entered school delayed, comparable to the German general population (2023: 6.2%)^24^, even though DSE is discussed frequently and emotionally with regard to preterm birth. DSE was associated with lower birth weight, lower gestational age and more perinatal complications. At the preschool follow-up (age five years) DSE was associated with lower IQ, more motor and behavioral problems. These difficulties^7^ may also appear in the school entry examination resulting in the decision for a DSE. The proportion of boys was significantly higher, compatible with other studies.^25^

Preterm infants in the DSE group consistently showed poorer outcomes in participation and in the F-words dimensions *Function* (especially SEN) and *Fitness*. No significant differences were observed in the dimensions *Friendship, Family* and *Fun*. However, after adjustment, no difference was found, suggesting a bias due to group distribution. Lower gestational age and the associated increased risk of complications have already been shown to result in more SEN.^26,27^ The results are consistent with other studies on academic outcomes of preterm^22,28^, extremely preterm children^29^ or term born children.^5^

We demonstrated that, in terms of *Future*, ASE supported timely transition to secondary school when accounting for all confounding factors. In contrast, ESE increased the likelihood of prolonged primary school attendance (i.e., repeating a school year). Other studies have shown that school problems can be avoided by correcting age for the preterm birth.^28,30^ Children who are one year older than their classmates appear to be more prone to behavioral problems, especially, if the child had to repeat a school year.^31^ Having to repeat a year appears to influence academic careers. In an US cohort study children who repeated a school year were less likely to graduate from high school.^32^ However, no long-term impact on term-born children’s well-being have been reported.^33^ Additionally, higher mortality rate and an increased risk of mental illness for children who had experienced school year retention were shown.^34,35^ Due to the high heterogeneity in school systems and the school entry policies nationally and internationally, results have to be compared cautiously. Within the DSE group, a larger proportion of VPT attended alternative or private schools compared to the other groups. This may be explained by the higher support needs of these children, which could also contribute to their later school entry.

Our results strongly support the consideration of age corrected for prematurity in the educational placement of preterm infants. When school entry is based on chronological age, VPT may be overestimated in their developmental readiness and thus enter school too early compared to their corrected age. Correcting for prematurity may help to avoid the increased risk of repeating a school year in primary school in the German school system. In terms of participation and quality of life, DSE does not appear to have any significant impact, up until the age ten years.

### Strength and limitations

Our systematic review was carried out in strict adherence to recommendations of the Cochrane methodological standards, thus, ensuring a high-quality literature search and synthesis. We conducted a comprehensive review of the literature using a broad search strategy. The inclusion criteria were predefined, and any deviations from the protocol, which was published prior to this review, were transparently documented. Two independent reviewers carried out the review process. The subsequent analysis provides data from a large German cohort of VPT. We were able to adjust analyses for many known confounders. Additionally, analysis of rare exposures was possible.

However, inherent limitations should be acknowledged. Although the overall sample was large, the DSE group was small, necessitating cautious interpretation. Randomized controlled trials for this topic are not feasible. The potential for selection bias is high, influenced by local politics and availability of adequate SEN-support locally in that year in Germany. Furthermore, residual confounding factors cannot be excluded, as the specific reasons for DSE were not captured. Academic outcomes could not be directly assessed. While the majority of the observed differences between the analyzed and overall cohort were clinically not significant, there has been a significant decrease in the proportion of mothers with a history of migration. This bias cannot be disregarded in a cohort study and must be considered when interpreting the results.

## Conclusion

The question of whether VPT should enter school delayed was deemed highly important during the update of the German follow-up care guideline. However, a systematic literature review revealed a substantial evidence gap, particularly regarding psychosocial outcomes. This study investigated the impact of DSE in VPT children and found no significant effect on participation or quality of life by the age of ten years. ASE, when based on corrected age for prematurity, was associated with a higher likelihood of completing primary school on time, without having to repeat a school year. For the German Guideline for Follow-up of Preterm-born Children the following recommendations were made by the group:

1. We recommend using the age corrected for prematurity to determine school entry.
2. We suggest not delaying school entry for preterm-born children.
3. We recommend to take reports from kindergarten and other professionals as well as environmental factors into account to decide about school entry for preterm-born children
4. In the event of a delayed school entry, we recommend that a corresponding support goal and implementation plan is available for the year before school enrollment.

## Data Availability

All data produced in the present study are available upon reasonable request to the authors

## Acknowledgments

We thank Judith Wohlleben, MD (University Hospital Würzburg, Department of Pediatrics), Marie Babel (University Hospital Würzburg, Department of Pediatrics) for their dedicated support during the screening process.

Furthermore, we express our sincere gratitude to Maria-Inti Metzendorf, DrPH, MSc (Institute of General Practice, Medical Faculty of the Heinrich-Heine-University Duesseldorf) and Heidrun Janka, MSc, Ma LIS (Institute of General Practice, Medical Faculty of the Heinrich-Heine-University Duesseldorf), for their expert assistance in developing and conducting the search strategy.

We also wish to thank Nina Gawehn, PhD (University of Applied Sciences Bochum), and Nele Stahlmann, MD (University Hospital Würzburg, Department of Pediatrics), for their valuable advice throughout the project.

In addition, we gratefully acknowledge the contributions of the following experts and representatives of scientific societies and professional associations: Maria Hitzschke, MD (Federal Association “Das frühgeborene Kind e.V.” (The Premature Child Association)), Gabriele Trost-Brinkhues, MD („Kindernetzwerk e.V.“ (Children’s Network Association); Professional Association of Pediatricians and Adolescent Physicians, BVKJ), Annette Mund, PhD („Kindernetzwerk e.V.“ (Children’s Network Association)), Harald Abele, MD (German Society of Gynecology and Obstetrics, DGGG), Anna-Katharina Rohlfs, MD (German Society of Oto-Rhino-Laryngology, Head and Neck Surgery, DGHNO-KHC; German Society of Phoniatrics and Pediatric Audiology, DGPP), Oda von Rahden, PhD (German Society of Midwifery Science, DGHWi), Lucas Wessel, MD (German Society of Pediatric Surgery, DGKCH), Susanne Bechtold-Dalla Pozza, MD (German Society of Pediatric Endocrinology and Diabetes, DGKED), Nelly Schulz-Weidner, MD (German Society of Pediatric Dentistry, DGKiZ), Bernd Wilken, MD (German Society of Pediatrics and Adolescent Medicine, DGKJ), Britta Hüning, MD (German Society of Pediatrics and Adolescent Medicine, DGKJ), Luise Poustka, MD (German Society of Child and Adolescent Psychiatry, Psychosomatics and Psychotherapy, DGKJP), Matthias Wagner, MD (German Society of Neuroradiology, DGNR), Christoph Härtel, MD (German Society for Pediatric Infectious Diseases, DGPI), Gernot Buheitel, MD (German Society for Pediatric Cardiology and Congenital Heart Defects, DGPK), Rolf Schlößer, MD (German Society for Perinatal Medicine, DGPM), Tim U. Krohne, MD (German Ophthalmological Society, DOG), Andreas Stahl, MD (German Ophthalmological Society, DOG), Regina Trollmann, MD (German-Speaking Society of Child Neurology, GNP), Claudia Roll, MD (Society for Neonatology and Pediatric Intensive Care Medicine, GNPI), Michael Zemlin, MD (Society for Neonatology and Pediatric Intensive Care Medicine, GNPI), Sandra Habbig, MD (German Society for Pediatric Nephrology, GPN), Stefanie Weber, MD (Society for Pediatric Nephrology, GPN), Anne Hilgendorff, MD (German Society for Pediatric Pneumology, GPP), Franz Wolfgang Hirsch, MD (German Society for Pediatric Radiology, GPR), Anja Helmers, MD (German-Speaking Association for Pediatric Orthopedics, VKO), Jana Zang, PhD (German Federal Association for Speech Therapy, dbl), Nicole Hübl, PhD (German Federal Association for Speech Therapy, dbl), Angela Ehlers, PhD (German Association for Special Education, vds), Kerstin Mieth, Diploma in Early Childhood Education for Children with Physical Disabilities (German Association for Interdisciplinary Early Intervention, VIFF) and Carolin Kraushaar, MSc (German Association for Physiotherapy, ZVK).

We would also like to thank Monika Nothacker, MD, MPH (Association of the Scientific Medical Societies – Institute for Medical Knowledge Management, AWMF-IMWi) for her invaluable support in providing methodological guidance throughout the development of the guideline.

## Conflict of Interest Disclosures (includes financial disclosures)

The authors have no conflicts of interest relevant to this article to disclose.

## Clinical Trial Registration (if any)

The cohort study was not registered. Data can be accessed via the authors upon request.

## Abbreviations

ASE: age-appropriate school entry
B: regression coefficient
BPD: bronchopulmonary dysplasia
BRIEF: Behavior Rating Inventory of Executive Function
CASP: Child and Adolescent Scale of Participation
CA: corrected age
CH: chronological age
CI: confidence interval
DSE: delayed school entry
ESE: early school entry
GNN: German Neonatal Network
ICF: International Classification of Functioning, Disability and Health
IQ: intelligence quotient
IVH: intraventricular hemorrhage
KOPKIJ: questionnaire for recognizing cognitive problems in children and adolescents
MABC: Movement Assessment Battery for Children
Mean: mean value
n: number
OR: odds ratio
POMP: Percent of Maximum Possible
sd: standard deviation
SDQ: Strengths and Difficulties Questionnaire
SEN: Special educational needs
VPT: very preterm children

## Article Summary

This investigates the associations of delayed school entry (DSE) in preterm born children and their academic performance, participation and behavioral outcomes.

## What’s Known on This Subject

Preterm infants are at an increased risk for neurological and behavioral problems. Consequently, some of these children do not meet school-readiness requirements and delaying school entry by a year is frequently discussed by parents as well as professionals.

## What This Study Adds

This study indicates that delaying school entry does not improve academic performance, behavior, reported quality of life or participation.

## Contributors Statement Page

*Dr. Anna Häfke and Antonia Schedewi conceptualized and designed the study, carried out the analysis, drafted the initial manuscript, and critically reviewed and revised the manuscript*.

*Antonia Schedewi, Carla Mehring, and Dr. Stephanie Weibel developed the protocol and supervised the systematic review process and critically reviewed and revised the manuscript*.

*Prof. Dr. Wolfgang Göpel supervised data collection and critically reviewed and revised the manuscript*.

*Prof. Dr. Juliane Spiegler designed the data collection instruments, supervised data collection, developed the protocol for the systematic review, and supervised the systematic review process, conceptualized and designed the study, coordinated, and critically reviewed and revised the manuscript for important intellectual content*.

*All authors approved the final manuscript as submitted and agree to be accountable for all aspects of the work*.

