## Supplemental file for "The Impact of Delayed School Entry on Developmental Outcomes in Very Preterm Infants"

| **Abbreviations** | **Description** |
|---|---|
| BASC | Behavior Assessment System for Children |
| BRIEF | Behaviour Rating Inventory of Executive Function |
| CASP | Child and Adolescent Scale of Participation |
| CBCL | Child Behavior Checklist |
| CHQ | Child Health Questionnaire |
| COPQ | Children's Occupational Performance Questionnaire |
| COSA | Child Occupational Self-Assessment |
| CPAS-P | Child-Parent Attachment Scale – Preschool version |
| CPQ | Child Perceptions Questionnaire |
| DRT-2 | "Diagnostischer Rechtschreibtest für 2. Klassen" (Diagnostic Spelling Test for second grade students) |
| GA | gestational age |
| GRADE | Grading of Recommendations, Assessment, Development and Evaluation |
| HRQoL | Health-Related Quality of Life |
| KOPKIJ | questionnaire for recognizing cognitive problems in children and adolescents |
| LBW | Low birth weight |
| PACS | Parental Account of Children's Symptoms |
| PedsQL™ | Pediatric Quality of Life Inventory™ |
| RCT | Randomized Controlled Trial |
| ROBINS-I | Risk Of Bias In Non-randomised Studies - of Interventions |
| RR | Risk ratio |
| SDQ | Strengths and Difficulties Questionnaire |

**Appendix 1.** Methods

The research question guiding this review was whether delayed school entry provides benefit or harm to preterm-born children in terms of educational success, quality of life, participation and behavior compared with age-appropriate school entry.

The protocol for this review was prospectively registered on PROSPERO (CRD42024620228).

Eligibility criteria

*Types of Studies*

While randomized controlled trials (RCTs) provide the highest level of evidence, such studies are unlikely to exist for this research question. Randomized allocation to delayed school entry would raise substantial ethical concerns, making RCTs largely unfeasible. Therefore, we primarily focused on observational studies such as prospective cohort studies, retrospective cohort studies, non-current cohort study, case control studies and cross-sectional study.

Eligible studies included published articles, registered trials (completed or ongoing), and completed but unpublished studies, which were categorized as "awaiting classification". Study authors were contacted for clarification where necessary.

Articles in another language than English or German were excluded due to resource limitations. Studies only available as conference abstracts were excluded because of limited methodological information.

*Participants/Population*

We included studies assessing:

- Children born preterm (<37 weeks gestational age (GA))
- Children born preterm AND with low birth weight (LBW <2500g)
- Children born preterm AND/OR with very low birth weight (VLBW <1500g)

        - Children born preterm AND/OR with extremely low birth weight (ELBW <1000g)

Studies including full-term children (>37 weeks GA) or children with LBW without detailed gestational information were excluded. Studies with mixed populations were only included if subgroup analyses of the defined population were performed.

*Intervention*

The intervention of interest was delayed school entry among preterm-born children in high-income countries. Delayed school entry was defined as school enrollment occurring after the corrected age of 6 years. Studies from countries including children who routinely enter school before the age of 6 years were excluded. Only studies conducted in high-income countries were included to ensure comparability with the German context.

*Comparator*

The comparator was age-appropriate school entry at 6 years of age.

*Outcomes*

We focused on a range of developmental outcomes, with particular interest in the following domains:

- School performance
    - Reading skills
    - Mathematical skills
    - Special educational needs
- Participation
    - International Classification of Functioning, Disability and Health measured with e.g. Child and Adolescent Scale of Participation (CASP), Child Perceptions Questionnaire (CPQ), Child Occupational Self-Assessment (COSA), Children's Occupational Performance

    Questionnaire (COPQ), Child-Parent Attachment Scale – Preschool version (CPAS-P), Parental Account of Children's Symptoms (PACS)

- Quality of life
    - Multidimensional tools e.g. Health-Related Quality of Life (HRQoL), Pediatric Quality of Life Inventory™ (PedsQL™), Child Health Questionnaire (CHQ), KidScreen8-52
    - Bullying scale
- Behavior
    - Externalizing problems: hyperactivity, oppositional behavior measured with e.g. Child Behavior Checklist (CBCL), Strengths and Difficulties Questionnaire (SDQ), Behavior Assessment System for Children (BASC)
    - Internalizing problems: anxiety, depression, somatization (CBCL, SDQ, BASC)
    - Adaptive skills: social skills, personal adjustment (CBCL, SDQ, BASC)

Additional outcomes were social life, atypicality and school problems.

Studies that did not report any of the prioritized outcomes were retained for documentation purposes. While their references were recorded, no data were extracted from these studies.

Search Methods

We searched the following platforms from inception until 13 November 2024: MEDLINE, PsycINFO, CINAHL, the Cochrane Central Register of Controlled Trials and the WHO International Clinical Trial Registry Platform (ICTRP). Search strategies are detailed in Appendix S2. Reference lists of identified primary studies and relevant systematic reviews were also screened.

Study selection was conducted according to our predefined eligibility criteria. First, two reviewers (AS, CM) independently screened titles and abstracts of identified records.

Second, full texts of potentially eligible studies were retrieved and assessed independently by both reviewers. Discrepancies at any stage were resolved through discussion or consultation with a third reviewer.

Screening was performed using Covidence.

Data Extraction

Data were extracted by two review authors (AS, CM) independently using Covidence.

Discrepancies were resolved through discussion.

For missing data, we contacted the authors of the concerned study.

Risk of Bias Assessment

Risk of bias was assessed using the "Risk Of Bias In Non-randomised Studies - of Interventions" (ROBINS-I)[1] tool. Two reviewers (AS, CM) independently evaluated each included study. Disagreements were resolved through discussion or another reviewer.

Studies were categorized according to ROBINS-I as having a low, moderate, serious or critical risk of bias, defined as follows:

- Low risk of bias - Few to none concerns regarding bias
- Moderate risk of bias – Some concerns regarding bias
- Serious risk of bias – Important methodological limitations
- Critical risk of bias – Very methodological flaws; grounds for

(continuous outcome) and Mantel-Haenszel method (binary outcomes) were prespecified.

Statistical heterogeneity was to be assessed using the $\chi^2$ test, the $I^2$ statistic, and – when at least 4 studies (≥200 participants) were available – 95% prediction intervals. Heterogeneity was defined a $P < 0.1$ for the $\chi^2$, $I^2 \geq 40\%$, or when the 95% prediction interval indicated a clinically different interpretation compared with the 95% CI. Subgroup analyses were planned according to gestational age and intervention characteristics. Sensitivity analyses based on the risk of bias and fixed-effect modelling were also prespecified. Certainty of the evidence was to be evaluated using the GRADE approach.

However, as only one eligible study was identified, no meta-analysis could be performed.

#### Certainty of the Evidence

The certainty of evidence was rated independently by two review authors (AS, CM) using the Grading of Recommendations, Assessment, Development and Evaluation tool (GRADE)[2]. End results can be high, moderate, low or very low evidence of certainty. Reasons for a downgrade of the certainty of evidence level were risk of bias, inconsistency, imprecision, indirectness, or publication bias. If disagreements occurred during the process these were resolved through discussion.

**Appendix 2. Search strategy**

**Database: Ovid MEDLINE(R) ALL <1946 to November 12, 2024>**

**1** exp Infant, Premature/ (68209)

**2** exp Infant, low birth weight/ (40158)

**3** Premature Birth/ (23659)

**4** ((premature or pre mature or preterm or pre term) adj3 (born or birth or infant* or neonat* or neo nat* or baby or babies or born? or newborn? or new born? or newly born? or child or children or adolescen*)).tw. (106132)

**5** ((neonatal or neo natal) adj3 (prematurity or pre maturity)).tw. (686)

**6** (low birth weight or low birthweight).tw. (41947)

**7** (preemie or premie or preemies or premies).tw. (231)

**8** or/1-7 (166638)

**9** Child Care/ or Infant Care/ (15682)

**10** (child care or child daycare or childcare or infant care or kindergarten* or kindergarden* or preschool* or pre-school* or nursery school* or day nurser* or pre-K or play school* or childhood education cent* or play group* or play center* or play centre* or daycare or day care or infant school* or non-parental care or non-relative care or nanny or nannies or babysitter* or child supervision or early childhood care or educational institution* or learning center*).tw. (77183)

**11** 9 or 10 (89235)

**12** (delay or delayed or postponed or late? or early or earlier or before the age or after the age).tw. (3409377)

**13** 8 and 11 and 12 (1080)

**14** Educational Measurement/ or Schools/ (97258)

**15** ((enroll* or admiss* or begin* or enter* or start* or attend* or entry) adj3 (school* or education*)).tw. (29961)

**16** 14 or 15 (121800)

**17** 8 and 16 (530)

**18** 13 or 17 (**1559**)

**CENTRAL via Cochrane Library**

**1.** [mh "Infant, Premature"]

**2.** [mh "Infant, low birth weight"]

**3**. [mh ^"Premature Birth"]

**4.** ((premature OR "pre mature" OR preterm OR "pre term") NEAR/3 (born OR birth OR infant* OR neonat* OR (neo NEXT nat*) OR baby OR babies OR born? OR newborn or newborns OR (new NEXT born) OR (new NEXT borns) OR (newly NEXT born) OR (newly NEXT borns) OR child OR children OR adolescen*)):ti,ab

**5.** ((neonatal OR "neo natal") NEAR/3 (prematurity OR "pre maturity")):ti,ab

**6.** ("low birth weight" OR "low birthweight"):ti,ab

**7.** (preemie OR premie OR preemies OR premies):ti,ab

**8.** #1 OR #2 OR #3 OR #4 OR #5 OR #6 OR #7

**9.** [mh ^"Child Care"] OR [mh ^"Infant Care"]

**10.** ("child care" OR "child daycare" OR childcare OR "infant care" OR kindergarten* OR kindergarden* OR preschool* OR pre-school* OR (nursery NEXT school*) OR (day NEXT nurser*) OR pre-K OR (play NEXT school*) OR ("childhood education" NEXT cent*) OR (play NEXT group*) OR (play NEXT center*) OR (play NEXT centre*) OR daycare OR "day care" OR (infant NEXT school*) OR "non-parental care" OR "non-relative care" OR nanny OR nannies OR babysitter* OR "child supervision" OR "early childhood care" OR (educational NEXT institution*) OR (learning NEXT center*)):ti,ab

**11.** #9 OR #10

**12.** (delay OR delayed OR postponed OR late OR late? OR early OR earlier OR "before the age" OR "after the age"):ti,ab

**13.** #8 AND #11 AND #12

**14.** [mh ^"Educational Measurement"]

**15.** [mh "Schools"]

**16.** ((enroll* OR admiss* OR begin* OR enter* OR start* OR attend* OR entry) NEAR/3 (school* OR education*)):ti,ab

**17.** #14 OR #15 OR #16

**18.** #8 AND #17

**19.** #13 OR #18

**20.** #19 in Trials

(**219**)

**APA PsycInfo <1806 to October 2024 Week 5>**

**1.** Premature Birth/
**2.** ((premature or pre mature or preterm or pre term) adj3 (born or birth or infant* or neonat* or neo nat* or baby or babies or born? or newborn? or new born? or newly born? or child or children or adolescen*)).tw.
**3.** ((neonatal or neo natal) adj3 (prematurity or pre maturity)).tw.
**4.** (low birth weight or low birthweight).tw.
**5.** (preemie or premie or preemies or premies).tw.
**6.** or/1-5
**7.** Child Day Care/ or Child Care/
**8.** (child care or child daycare or childcare or infant care or kindergarten* or kindergarden* or preschool* or pre-school* or nursery school* or day nurser* or pre-K or play school* or childhood education cent* or play group* or play center* or play centre* or daycare or day care or infant school* or non-parental care or non-relative care or nanny or nannies or babysitter* or child supervision or early childhood care or educational institution* or learning center*).tw.
**9.** 7 or 8
**10.** (delay or delayed or postponed or late? or early or earlier or before the age or after the age).tw.
**11.** 6 and 9 and 10
**12.** exp Educational Measurement/ or Student Learning Outcomes/ or Schools/
**13.** ((enroll* or admiss* or begin* or enter* or start* or attend* or entry) adj3 (school* or education*)).tw.
**14.** 12 or 13
**15.** 6 and 14
**16.** 11 or 15 **(531)**

**CINAHL (EBSCO)**

1. (MH "Childbirth, Premature") OR (MH "Infant, Premature")
2. TI ((premature OR  "pre mature" OR preterm OR "pre term") N3 (born OR birth OR infant* OR neonat* OR "neo nat*" OR baby OR babies OR born# OR newborn# OR "new born#" OR "newly born#" OR child OR children OR adolescen*)) OR AB ((premature OR "pre mature" OR preterm OR "pre term") N3 (born OR birth OR infant* OR neonat* OR "neo nat*" OR baby OR babies OR born# OR newborn# OR "new born#" OR "newly born#" OR child OR children OR adolescen*))
3. TI ((neonatal OR "neo natal") N3 (prematurity OR "pre maturity")) OR AB ((neonatal OR "neo natal") N3 (prematurity OR "pre maturity"))
4. TI ("low birth weight" OR "low birthweight") OR AB ("low birth weight" OR "low birthweight")
5. TI (preemie OR premie OR preemies OR premies) OR AB (preemie OR premie OR preemies OR premies)
6. S1 OR S2 OR S3 OR S4 OR S5
7. (MH "Child Care") OR (MH "Child Day Care")
8. TI ("child care" OR "child daycare" OR childcare OR "infant care" OR kindergarten* OR kindergarden* OR preschool* OR "pre-school*" OR "nursery school*" OR "day nurser*" OR "pre-K" OR "play school*" OR  "childhood education cent*" OR "play group*" OR "play center*" OR "play centre*" OR daycare OR "day care" OR "infant school*" OR "non-parental care" OR "non-relative care" OR nanny OR nannies OR babysitter* OR "child supervision" OR "early childhood care" OR "educational institution*" OR "learning center*") OR AB ("child care" OR "child daycare" OR childcare OR "infant care" OR kindergarten* OR kindergarden* OR preschool* OR "pre-school*" OR "nursery school*" OR "day nurser*" OR "pre-K" OR "play school*" OR  "childhood education cent*" OR "play group*" OR "play center*" OR "play centre*" OR daycare OR "day care" OR "infant school*" OR "non-parental care" OR "non-relative care" OR nanny OR nannies OR babysitter* OR "child supervision" OR "early childhood care" OR "educational institution*" OR "learning center*")
9. S7 OR S8
10. TI (delay OR delayed OR postponed OR late# OR early OR earlier OR "before the age" OR "after the age") OR AB (delay OR delayed OR postponed OR late# OR early OR earlier OR "before the age" OR "after the age")
11. S6 AND S9 AND S10
12. (MH "Educational Measurement")
13. (MH "Schools")
14. TI ((enroll* OR admiss* OR begin* OR enter* OR start* OR attend* OR entry) N3 (school* OR education*)) OR AB ((enroll* OR admiss* OR begin* OR enter* OR start* OR attend* OR entry) N3 (school* OR education*))
15. S12 OR S13 OR S14
16. S6 AND S14
17. S11 OR S15 **(557)**

**WHO ICTRP Database**

((prematur* OR "pre mature" OR preterm OR "pre term" OR "low birth weight" OR "low birthweight") AND (born OR borns OR birth OR infant* OR neonat* OR "neo nat*" OR baby OR babies OR newborn* child OR children)) AND ((("child care" OR childcare OR daycare OR "day care" OR "infant care" OR kindergarten* OR kindergarden* OR preschool* OR "pre school*" OR "nursery school*" OR "day nurser*" OR "play school*" OR "childhood education cent*" OR "play center*" OR "play centre*" OR "infant school*" OR "non parental care" OR "non relative care" OR nann* OR babysitter* OR "child supervision" OR "childhood care" OR "educational institution*" OR "learning center*") AND (delay OR delayed OR postponed OR late OR later OR early OR earlier)) OR ((enroll* OR admiss* OR begin* OR enter* OR start* OR attend* OR entry) AND (school* OR education*)))

**(88)**

### Appendix 3. Results

Search

The search strategy (Appendix S2) identified 2,866 records in databases and 88 records in trial registers. After removing 874 records, 2,080 studies were screened. During the title and abstracts screening 1,898 studies were excluded. For one of the 182 remaining studies we could not retrieve a full-text article. 181 remained for the full-text screening. 180 studies were excluded. One study was included (Figure S1).

An overview of all included and excluded studies along with their reasons for exclusion is provided in Table S4.

Characteristics of included studies

One study was included, comprising 340 preterm-born children aged 6 to 8 years. The study was based on data from the Bavarian Longitudinal Study conducted in Bavaria, Germany, and published in 2015[3].

It investigated educational and attentional outcomes among preterm-born children during their first and second years of formal schooling, taking age at school entry into account. Children with delayed school entry formed the intervention group, while those with age-appropriate school entry served as the control group.

At the end of the first school year, teachers rated the children's abilities in writing, reading, mathematics, and attention. In the second school year, standardized assessments were administered:

- Writing was assessed using the "Diagnostischer Rechtschreibtest für 2. Klassen" (DRT-2, Diagnostic Spelling Test for second grade students)
- Reading was measured with the Zürich Reading Test and a pseudoword reading test
- Mathematics was evaluated using a standardized math test
- Attention regulation during standardized testing was rated using the task orientation subscale of the Testers Rating of Child Behavior

Additional information is provided in Table S1 and S2.

Risk of Bias

ROBINS-I were used to assess the Risk of Bias of the outcomes of the remaining study (Table S5).

Outcomes based on teacher ratings were judged to be at serious risk of bias and were excluded from further consideration.

Outcomes based on standardized assessments were rated as having a low risk of bias.

Effects of interventions

*School performance*

School performance was assessed at the age of 8 years across three domains: reading, writing and mathematics. In all areas, children with delayed school entry performed worse on standardized tests than those with age-appropriate school entry (Table S5). Standardized scores (mean=100, standard deviation=15) were obtained directly from the original authors, as the published article reported only z-scores.

However, all assessments were conducted at the same time point, meaning that children with delayed school entry had received approximately 8 months less formal instruction. This difference in schooling duration was accounted for in the analysis. The expected improvement in test performance over 8 months, based on the progressions observed in the age-appropriate school entry group, was projected onto the delayed school entry group. Even after adjusting for this estimated learning gain, children with delayed school entry continued to score lower across all domains (graphically extracted from Figure S2; 30.2% (reading), 11.3% (writing), 47.2% (mathematics) were predicted to reach the performance level of age-appropriate school entry children).

The certainty of evidence for all three domains was rated as very low due to imprecision (Table 2). The evidence on the effect of delayed school entry on educational performance remained unclear.

*Attention*

Attention was evaluated as an additional developmental domain. As with the academic outcomes, children with delayed school entry scored lower as children with age-appropriate school entry (Table S5). Similar to school performances, this finding persisted after adjusting for differences in schooling duration (graphically extracted from Figure S2; 28.3% were predicted to reach the attention-performance level of age-appropriate school entry children).

The certainty of evidence was rated as very low due to imprecision. The evidence on the effect of delayed school entry on behavioral problems remained unclear.

**Supplemental-Table S 1.** Summary of included studies

| **Study reference** | **Country** | **Popula-tion** | **Randomized Patients** | | **GA in weeks** | | **Age at assessment in years** | **Intervention** | **Comparator** | **Outcome** |
|---|---|---|---|---|---|---|---|---|---|---|
| | | | **CG** | **IG** | **CG** | **IG** | | | | |
| **Jaekel 2015** | Germany | GA < 37 weeks | n = 287 | n = 53 | < 37 | < 37 | 8 | delayed school entry | age-appropriate school entry | School performance<br>mathematical skills (standardized test)<br>reading skills (Zürich reading test and pseudoword reading test)<br>writing skills (DRT-2)<br>Behavior<br>attention (task orientation subscale of the Tester's Rating of Child Behavior) |

CG comparator group, DRT "Diagnostischer Rechtschreibtest für 2. Klassen" (Diagnostic Spelling Test for second grade students), GA gestational age, IG intervention group

**Supplemental-Table S 2.** Detailed characteristics of included studies

| **Study ID** | **Jaekel 2015** |
|---|---|
| **General information** | |
| Title | Delayed school entry and academic performance: a natural experiment |
| Primary study report | DOI: 10.1111/dmcn.12713 |
| Lead author contact details | Dieter Wolke at Department of Psychology and Warwick Medical School, University of Warwick, Coventry, CV47AL, UK.<br> |
| Possible conflicts of interest for study authors | The authors have stated that they had no interests that might be perceived as posing a conflict or bias. |
| Start date | xx/xx/1990 |
| End date | xx/xx/1993 |
| Study funding sources | non-industrial<br>This study was supported by grant EDU/40442 from the Nuffield Foundation. The Nuffield Foundation was not involved at any stage in the drafting of the manuscript. The contents are solely the responsibility of the authors and do not necessarily represent the official views of the Nuffield Foundation. There are no financial relationships with any organizations that might have an interest in the submitted work in the previous 3 years. |
| Country | Germany |
| Status of study | completed and published as journal publication or preprint |
| Journal | Developmental Medicine & Child Neurology |
| **Methods** | |
| Aim of study | The aim of this study was to investigate the effect of delayed school entry on children’s academic performance. |
| Study design | Prospective Cohort study |
| Additional study reports available | Published as journal publication or preprint |
| Setting | Bavarian Longitudinal Study |
| **Participants** | |
| Inclusion criteria | Children born in 1985 and 1986 within a geographically defined area of Southern Bavaria (Germany) who required admission to a children’s hospital within the first 10 days of life. (n=7505; 10.6% of all live births). Additionally, 916 healthy term-born control infants were identified at birth from the same hospitals in Bavaria. Of the initial 8421 children, 1316 survivors stratified by |

| | |
|---|---|
| | sex, socio-economic status, and degree of neonatal risk were assessed at 6 and 8 years of age. |
| Exclusion criteria | Of these, 118 children were excluded who were either (1) born at more than 41 weeks' gestation, (2) entered school 1 year early (in Germany, parents can ask to have their child tested for early school entry), or (3) were enrolled directly in a school for children with special needs (i.e. teacher ratings would not be comparable). In addition, 199 children had incomplete information on baseline characteristics and could not be included in PSM. |
| Baseline population characteristics | |
| Preterm as… | subgroup |
| Gestational age | <37 SSW |
| Birth weight | Not reported |
| Total number of participants - overall | 340 |
| Total number of participants - intervention | 53 |
| Total number of participants - comparator | 287 |
| Sociodemographic characteristics of parents | Socioeconomic status<br>Intervention group:<br>Low: 17 (32%); middle: 23 (44%); high: 13 (24%)<br>Comparator<br>Low: 101 (35%); middle: 123 (43%); high: 63 (22%) |
| **Intervention** | |
| Type of intervention | Delayed school entry |
| Subgroup analysis | none |
| **Comparator** | |
| Description of comparator | Age-appropriate school entry |
| **Outcomes** | |
| Educational success/ school performance | teacher ratings<br>children's standardized test scores (mathematics test, Zürich Reading Test and pseudoword reading test, diagnostic spelling test: writing and spelling)<br>(7-12 years) |
| Behavior | attention regulation: task orientation sub-scale of the Tester's Rating of Child Behaviour<br>(7-12 years) |

**Supplemental-Table S 3.** List of all included and excluded studies

| Study ID | Title | DOI | Reason for exclusion |
|---|---|---|---|
| Jaekel 2015 | Delayed school entry and academic performance: a natural experiment | https://dx.doi.org/10.1111/dmcn.12713 | |
| Alenius 2023 | School grades and educational attainments of adolescents and young adults born preterm | https://dx.doi.org/10.1038/s41598-022-27295-4 | Wrong intervention |
| Andrews 1995 | Prediction of special education placement from birth certificate data | | Wrong patient population |
| Bandyopadhyay 2023 | Factors associated with low school readiness, a linked health and education data study in Wales, UK | https://dx.doi.org/10.1371/journal.pone.0273596 | Wrong intervention |
| Bantel 2019 | [Factors Associated with Behavioral Problems in Pre-School Age: Secondary Data Analysis of Routine Examination at School Enrolment 2010/2014 in the Hannover Region] | https://dx.doi.org/10.1055/a-0594-2373 | Wrong study design |
| Barile 2024 | The Prematurity Paradox: Reevaluating the Kindergarten Readiness of Former Preterm Infants | https://dx.doi.org/10.1542/peds.2023-063801 | Wrong study design |
| Bauer 2010 | Kindergarten readiness after prematurity: Integrating health, development, and behavioral functioning to optimize educational outcomes of vulnerable children | https://dx.doi.org/10.1002/ddrr.126 | Wrong study design |
| Beauregard 2018 | Does Socioeconomic Status Modify the Association Between Preterm Birth and Children's Early Cognitive Ability and Kindergarten Academic Achievement in the United States? | https://dx.doi.org/10.1093/aje/kwy068 | Wrong intervention |
| Berbis 2012 | Quality of life of early school-age French children born preterm: a cohort study | https://dx.doi.org/10.1016/j.ejogrb.2012.02.006 | Wrong intervention |
| Berge 2023 | [Effects of High Birth Weight on the Development of Preschoolers] | https://dx.doi.org/10.1055/a-2160-0584 | Wrong intervention |
| Berry 2008 | The influence of specific aspects of preschool children's language on growth in reading comprehension across elementary school | | Wrong intervention |
| Bettge 2014 | Birth weight and special educational needs: results of a population-based study in Berlin | https://dx.doi.org/10.3238/arztebl.2014.0337 | Wrong intervention |

| | | | |
|---|---|---|---|
| Blair 2001 | The early identification of risk for grade retention among African American children at risk for school difficulty | https://dx.doi.org/10.1207/S1532480XADS0501_4 | Wrong intervention |
| Bohnert 2008 | Stability of psychiatric outcomes of low birth weight: a longitudinal investigation | https://dx.doi.org/10.1001/archpsyc.65.9.1080 | Wrong intervention |
| Borden 1997 | Early arrival | | full text unavailable |
| Breslau 2006 | Low birthweight and social disadvantage: Tracking their relationship with children's IQ during the period of school attendance | https://dx.doi.org/10.1016/j.intell.2005.10.003 | Wrong intervention |
| Breslau 2001 | Academic achievement of low birthweight children at age 11: the role of cognitive abilities at school entry | https://dx.doi.org/10.1023/a:1010396027299 | Wrong intervention |
| Breslau 2004 | The lingering academic deficits of low birth weight children | https://dx.doi.org/10.1542/peds.2004-0069 | Wrong intervention |
| Brosch-Fohraheim 2019 | The influence of preterm birth on expressive vocabulary at the age of 36 to 41 months | https://dx.doi.org/10.1097/MD.0000000000014404 | Wrong intervention |
| Burstein 2024 | Neonatal care and developmental outcomes following preterm birth: A systematic review and meta-analysis | https://dx.doi.org/10.1037/dev0001844 | Wrong intervention |
| Candel-Pau 2016 | Neurodevelopment in preterm infants with and without placenta-related intrauterine growth restriction and its relation to perinatal and postnatal factors | https://dx.doi.org/10.3109/14767058.2015.1081893 | Wrong intervention |
| Carter 2017 | Language abilities as a framework for understanding emerging cognition and social competencies after late, moderate, and very preterm birth | https://dx.doi.org/10.1016/j.jpeds.2016.10.077 | Wrong study design |
| Carvalho 2001 | Developmental history and behavior of pre-term and low birth-weight children (<1,500 g) | https://dx.doi.org/10.1590/S0102-79722001000100002 | Wrong language |
| Charkaluk 2011 | Very preterm children free of disability or delay at age 2: predictors of schooling at age 8: a population-based longitudinal study | https://dx.doi.org/10.1016/j.earlhumdev.2011.01.033 | Wrong intervention |
| Chaudhari 2005 | Biology versus environment in low birth weight children | | Wrong patient population |
| Chen 2014 | Prematurity and school readiness in a nationally representative sample of Australian children: | https://dx.doi.org/10.1016/j.earlhumdev.2013.09.015 | Wrong intervention |

| | | | |
|---|---|---|---|
| | does typically occurring preschool moderate the relationship? | | |
| Chyi 2008 | School outcomes of late preterm infants: special needs and challenges for infants born at 32 to 36 weeks gestation | https://dx.doi.org/10.1016/j.jpeds.2008.01.027 | Wrong intervention |
| Corrigan 1967 | The influence of prematurity on school performance | | Wrong intervention |
| Crockett 2022 | Education Outcomes of Children Born Late Preterm: A Retrospective Whole-Population Cohort Study | https://dx.doi.org/10.1007/s10995-022-03403-8 | Wrong intervention |
| De Hirsch 1966 | Comparisons between prematurely and maturely born children at three age levels | https://dx.doi.org/10.1111/j.1939-0025.1966.tb02313.x | Wrong intervention |
| De Leeuw 2024 | Socio-familial environment influence on cognitive and language development in very preterm children | https://dx.doi.org/10.1111/cch.13239 | Wrong intervention |
| De Rose 2013 | Perceptual-motor abilities in pre-school preterm children | https://dx.doi.org/10.1016/j.earlhumdev.2013.07.001 | Wrong intervention |
| DeBattista 2015 | Factors influencing early trajectories of adaptive behavior in children born prematurely | | Wrong intervention |
| Dempsey 2015 | School-aged children born preterm: Review of functioning across multiple domains and guidelines for assessment | https://dx.doi.org/10.1080/1754730X.2014.978117 | Wrong study design |
| Doyle 2021 | School-aged neurodevelopmental outcomes for children born extremely preterm | https://dx.doi.org/10.1136/archdischild-2021-321668 | Wrong study design |
| Drillien 1980 | Low-birthweight children at early school-age: a longitudinal study | https://dx.doi.org/10.1111/j.1469-8749.1980.tb04303.x | Wrong intervention |
| Drummond 2019 | Educational and health outcomes associated with bronchopulmonary dysplasia in 15-year-olds born preterm | https://dx.doi.org/10.1371/journal.pone.0222286 | Wrong intervention |
| Druschke 2020 | Individual-Level Linkage of Primary and Secondary Data from Three Sources for Comprehensive Analyses of Low Birthweight Effects | https://dx.doi.org/10.1055/a-1082-0740 | Wrong study design |
| Durlak 2016 | Relationship between Proton Magnetic Resonance Spectroscopy of Frontoinsular Gray Matter and Neurodevelopmental Outcomes in | https://dx.doi.org/10.1371/journal.pone.0156064 | Wrong intervention |

| | | | |
|---|---|---|---|
| | Very Low Birth Weight Children at the Age of 4 | | |
| Elgen 2002 | Low birthweight children: coping in school? | https://dx.doi.org/10.1080/080352502760148676 | Wrong intervention |
| Elgen 2012 | Mental health at 5 years among children born extremely preterm: a national population-based study | https://dx.doi.org/10.1007/s00787-012-0298-1 | Wrong intervention |
| Erdei 2020 | Predicting School-Aged Cognitive Impairment in Children Born Very Preterm | https://dx.doi.org/10.1542/peds.2019-1982 | Wrong intervention |
| Fevang 2017 | Mental health assessed by the Strengths and Difficulties Questionnaire for children born extremely preterm without severe disabilities at 11 years of age: a Norwegian, national population-based study | 10.1007/s00787-017-1007-x | Wrong intervention |
| Finnstrom 2000 | [School maladjustment common among children with very low birth weight Special attention and support are required during school start] | | Wrong language |
| Forsyth 2016 | A simple screen performed at school entry can predict academic under-achievement at age 7 in children born very preterm | https://dx.doi.org/10.1111/jpc.13361 | Wrong intervention |
| Foster-Cohen 2010 | High prevalence/low severity language delay in preschool children born very preterm | https://dx.doi.org/10.1097/DBP.0b013e3181e5ab7e | Wrong intervention |
| Frisk 1963 | Small Prematures at 6-7 Years of Age. Ii. School Maturity | | Wrong intervention |
| Gaddlin 2008 | "Academic achievement, behavioural outcomes and MRI findings at 15 years of age in very low birthweight children:" Corrigendum | https://dx.doi.org/10.1111/j.1651-2227.2008.01138_2.x | Wrong intervention |
| Gaddlin 2011 | Follow-up studies of very low birthweight children in Sweden | https://dx.doi.org/10.1111/j.1651-2227.2011.02288.x | Wrong study design |
| Gaddlin 2008 | Academic achievement, behavioural outcomes and MRI findings at 15 years of age in very low birthweight children | https://dx.doi.org/10.1111/j.1651-2227.2008.00925.x | Wrong intervention |
| Galan-Megias 2021 | Interaction of Impulsivity, Attention, and Intelligence in Early Adolescents Born Preterm without Sequelae | https://dx.doi.org/10.3390/ijerph18179043 | Wrong intervention |
| Gansaonre 2023 | Birthweight, gestational age, and early school trajectory | https://dx.doi.org/10.1186/s12889-023-15913-3 | Wrong study design |
| Garfield 2017 | Educational Performance of Children Born | https://dx.doi.org/10.1001/jamapediatrics.2017.1020 | Wrong intervention |

| | | | |
|---|---|---|---|
| | Prematurely | | |
| Georgsdottir 2012 | Disabilities and health of extremely low-birthweight teenagers: a population-based study | https://dx.doi.org/10.1111/j.1651-2227.2011.02576.x | Wrong intervention |
| Goldenberg 1996 | Pregnancy outcome and intelligence at age five years | https://dx.doi.org/10.1016/s0002-9378(96)70099-6 | Wrong intervention |
| Guarini 2014 | Basic numerical processes in very preterm children: a critical transition from preschool to school age | https://dx.doi.org/10.1016/j.earlhumdev.2013.11.003 | Wrong intervention |
| Gurka 2010 | Long-term cognition, achievement, socioemotional, and behavioral development of healthy late-preterm infants | https://dx.doi.org/10.1001/archpediatrics.2010.83 | Wrong intervention |
| Hall 1995 | School attainment, cognitive ability and motor function in a total Scottish very-low-birthweight population at eight years: a controlled study | https://dx.doi.org/10.1111/j.1469-8749.1995.tb11965.x | Wrong intervention |
| Hanke 2003 | Preschool development of very low birth weight children born 1994-1995 | https://dx.doi.org/10.1007/s00431-002-1127-1 | Wrong intervention |
| Hayes 2005 | Predictors of grade retention and special education placement: Implications for improving educational outcomes in the state of Florida | | Wrong intervention |
| Heuser 2018 | Origins and Predictors of Friendships in 6- to 8-Year-Old Children Born at Neonatal Risk | https://dx.doi.org/10.1016/j.jpeds.2017.09.072 | Wrong intervention |
| Hogan 2000 | Family factors and social support in the developmental outcomes of very low-birth weight children | https://dx.doi.org/10.1016/s0095-5108(05)70030-0 | Wrong intervention |
| Huning 2021 | Preterm birth and long-term consequences up to school age. Implications for follow-up services and schooling | https://dx.doi.org/10.1026/0942-5403/a000326 | Wrong study design |
| Jain 2008 | School outcome in late preterm infants: a cause for concern | https://dx.doi.org/10.1016/j.jpeds.2008.03.001 | Wrong intervention |
| Jansen 2020 | Classroom-evaluated school performance at nine years of age after very preterm birth | https://dx.doi.org/10.1016/j.earlhumdev.2019.104834 | Wrong intervention |
| Johnson 2009 | Academic attainment and special educational needs in extremely preterm children at 11 years of age: the EPICure study | https://dx.doi.org/10.1136/adc.2008.152793 | Wrong intervention |
| Kelly 2023 | A test of differential susceptibility in behavior trajectories of preterm infants from preschool to | https://dx.doi.org/10.1002/nur.22275 | Wrong intervention |

| | | | |
|---|---|---|---|
| | adulthood | | |
| Kiechl-Kohlendorfer 2013 | Early risk predictors for impaired numerical skills in 5-year-old children born before 32 weeks of gestation | https://dx.doi.org/10.1111/apa.12036 | Wrong intervention |
| Kitchen 1989 | Selective improvement in cognitive test scores of extremely low birthweight infants aged between 2 and 5 years | https://dx.doi.org/10.1111/j.1440-1754.1989.tb01479.x | Wrong intervention |
| Kitchen 1980 | A longitudinal study of very low-birthweight infants. IV: An overview of performance at eight years of age | https://dx.doi.org/10.1111/j.1469-8749.1980.tb04326.x | Wrong intervention |
| Klein 1985 | Preschool performance of children with normal intelligence who were very low-birth-weight infants | | Wrong intervention |
| Klimek 2018 | Temperament traits in 4-year-old children born prematurely - may they suggest a threat for mental functioning? | https://dx.doi.org/10.12740/PP/OnlineFirst/66229 | Wrong language |
| Koc 2016 | School Performance and Neurodevelopment of Very Low Birth Weight Preterm Infants: First Report From Turkey | https://dx.doi.org/10.1177/0883073815587028 | Wrong intervention |
| Kull 2016 | Early physical health problems as developmental liabilities for school readiness: Associations with early learning contexts and family socioeconomic status | | Wrong intervention |
| Largo 1990 | Intellectual outcome, speech and school performance in high risk preterm children with birth weight appropriate for gestational age | https://dx.doi.org/10.1007/BF02072071 | Wrong intervention |
| Larroque 2011 | Special care and school difficulties in 8-year-old very preterm children: the Epipage cohort study | https://dx.doi.org/10.1371/journal.pone.0021361 | Wrong intervention |
| Leversen 2012 | Prediction of outcome at 5 years from assessments at 2 years among extremely preterm children: a Norwegian national cohort study | https://dx.doi.org/10.1111/j.1651-2227.2011.02504.x | Wrong intervention |
| Liebhardt 2000 | Visual-motor function of very low birth weight and full-term children at 3 1/2 to 4 years of age | https://dx.doi.org/10.1016/s0378-3782(99)00056-0 | Wrong intervention |
| Liu 2020 | [Neurobehavioral development of 25 254 children with different gestational ages at birth in three cities of China] | | Wrong language |

| | | | |
|---|---|---|---|
| Lock 2024 | Academic performance in moderately and late preterm children in the United States: are they catching up? | https://dx.doi.org/10.1038/s41372-024-01938-y | Wrong intervention |
| Machova 1984 | Prematureness as a risk factor in school work | | Wrong language |
| Majnemer 2017 | Educational and rehabilitation service utilization in adolescents born preterm or with a congenital heart defect and at high risk for disability | https://dx.doi.org/10.1111/dmcn.13520 | Wrong intervention |
| Mangin 2017 | Cognitive Development Trajectories of Very Preterm and Typically Developing Children | https://dx.doi.org/10.1111/cdev.12585 | Wrong intervention |
| Markkula 2024 | Interventions and their efficacy in supporting language development among preterm children aged 0-3 years - A systematic review | https://dx.doi.org/10.1016/j.earlhumdev.2024.106057 | Wrong study design |
| Martin 2007 | The effects of prematurity and parent stress on preschoolers' behavior and temperament as rated by parents and teachers | | Wrong intervention |
| Maupin 2014 | Differential effects of parenting in preterm and full-term children on developmental outcomes | https://dx.doi.org/10.1016/j.earlhumdev.2014.08.014 | Wrong intervention |
| McGowan 2022 | Neurodevelopmental Outcomes Associated with Prematurity from Infancy Through Early School Age: A Literature Review | 10.62116/pnj.2022.48.5.223 | Wrong study design |
| McGrath 2000 | Longitudinal neurologic follow-up in neonatal intensive care unit survivors with various neonatal morbidities | https://dx.doi.org/10.1542/peds.106.6.1397 | Wrong intervention |
| Meio 2004 | [Pre-school cognitive development of very low birth weight preterm children] | | Wrong intervention |
| Mercier 2022 | Neurodevelopment at seven years and parents' feelings of prematurely born children | https://dx.doi.org/10.3389/fped.2022.1004785 | Wrong intervention |
| Miller 2018 | Executive functioning in low birth weight children entering kindergarten | https://dx.doi.org/10.1038/jp.2017.147 | Wrong intervention |
| Monset-Couchard 1996 | Mid- and long-term outcome of 89 premature infants weighing less than 1,000 g at birth, all appropriate for gestational age | https://dx.doi.org/10.1159/000244384 | Wrong intervention |
| Monset-Couchard 2002 | Mid- and long-term outcome of 166 premature infants weighing less than 1,000 g at birth, all small for gestational age | https://dx.doi.org/10.1159/000056755 | Wrong study design |
| Morse 2009 | Early school-age outcomes of late preterm | https://dx.doi.org/10.1542/peds.2008-1405 | Wrong intervention |

| | | | |
|---|---|---|---|
| | infants | | |
| Msall 2014 | Commentary on "Kindergarten classroom functioning of extremely preterm/extremely low birth weight children" or "Leaving no child behind: promoting educational success for preterm survivors" | https://dx.doi.org/10.1016/j.earlhumdev.2014.10.002 | Wrong intervention |
| Msall 2002 | Measuring functional outcomes after prematurity: developmental impact of very low birth weight and extremely low birth weight status on childhood disability | https://dx.doi.org/10.1002/mrdd.10046 | Wrong study design |
| Munck 2012 | Stability of cognitive outcome from 2 to 5 years of age in very low birth weight children | https://dx.doi.org/10.1542/peds.2011-1566 | Wrong intervention |
| Nagy 2018 | [Follow-up study of extremely low birth weight preterm infants to preschool age in the light of perinatal complications] | https://dx.doi.org/10.1556/650.2018.31199 | Wrong language |
| O'Callaghan 1996 | School performance of ELBW children: a controlled study | https://dx.doi.org/10.1111/j.1469-8749.1996.tb15048.x | Wrong intervention |
| O'Meagher 2017 | Risk factors for executive function difficulties in preschool and early school-age preterm children | https://dx.doi.org/10.1111/apa.13915 | Wrong intervention |
| Odberg 2011 | Low birth weight young adults: quality of life, academic achievements and social functioning | https://dx.doi.org/10.1111/j.1651-2227.2010.02096.x | Wrong patient population |
| Pacheco 2018 | Low weight at birth and unfavorable socioeconomic background: Effects on language and fluid intelligence | https://dx.doi.org/10.1080/17450128.2018.1524613 | Wrong patient population |
| Patrianakos-Hoobler 2009 | Risk factors affecting school readiness in premature infants with respiratory distress syndrome | https://dx.doi.org/10.1542/peds.2008-1771 | Wrong intervention |
| Peng 2005 | Outcome of low birthweight in China: a 16-year longitudinal study | https://dx.doi.org/10.1111/j.1651-2227.2005.tb01999.x | Wrong intervention |
| Perez-Roche 2016 | Effect of prematurity and low birth weight in visual abilities and school performance | https://dx.doi.org/10.1016/j.ridd.2016.10.002 | Wrong intervention |
| Pfeiffer 1990 | Outcome for preschoolers of very low birthweight: sociocultural and environmental influences | https://dx.doi.org/10.2466/pms.1990.70.3c.1367 | Wrong intervention |
| Pharoah 1994 | Clinical and subclinical deficits at 8 years in a geographically defined cohort of low birthweight | https://dx.doi.org/10.1136/adc.70.4.264 | Wrong intervention |

| | | | |
|---|---|---|---|
| | infants | | |
| Ramey 1999 | Prevention of intellectual disabilities: Early interventions to improve cognitive development | | Wrong study design |
| Rantakallio 1985 | Prognosis for low-birthweight infants up to the age of 14: a population study | https://dx.doi.org/10.1111/j.1469-8749.1985.tb14138.x | Wrong intervention |
| Redemann 2021 | [Association between Socioeconomic Status and Developmental Status: Data Linkage of Results of the Daycare and School Entry Health Examinations of a Saxon Birth Cohort Study] | https://dx.doi.org/10.1055/a-1327-2463 | Wrong intervention |
| Reid 2019 | A Population-Based Study of School Readiness Determinants in a Large Urban Public School District | https://dx.doi.org/10.1007/s10995-018-2666-z | Wrong intervention |
| Reuner 2009 | Long-term development of low-risk low birth weight preterm born infants: neurodevelopmental aspects from childhood to late adolescence | https://dx.doi.org/10.1016/j.earlhumdev.2009.01.007 | Wrong patient population |
| Riser 2019 | Native American children and school readiness: A nationally representative study of individual and cumulative risks | https://dx.doi.org/10.1016/j.childyouth.2019.104496 | Wrong intervention |
| Saigal 2003 | School-age outcomes in children who were extremely low birth weight from four international population-based cohorts | https://dx.doi.org/10.1542/peds.112.4.943 | Wrong intervention |
| Salm 2008 | Child health disparities, socio-economic status, and school enrollment decisions: evidence from German elementary school entrance exams | | Wrong patient population |
| Sansavini 2010 | Does preterm birth increase a child's risk for language impairment? | https://dx.doi.org/10.1016/j.earlhumdev.2010.08.014 | Wrong intervention |
| Santos 2008 | Determinants of cognitive function in childhood: a cohort study in a middle income context | https://dx.doi.org/10.1186/1471-2458-8-202 | Wrong patient population |
| Sawyer 2021 | Self-regulation task in young school age children born preterm: Correlation with early academic achievement | https://dx.doi.org/10.1016/j.earlhumdev.2021.105362 | Wrong intervention |
| Schaap 1999 | School performance and behaviour in extremely preterm growth-retarded infants | https://dx.doi.org/10.1016/s0301-2115(99)00041-x | Wrong intervention |
| Schlosser 2008 | [Impact of preterm infants of less than 30 weeks gestation on the prevalence of special education | https://dx.doi.org/10.1055/s-2007-984375 | Wrong intervention |

| | | | |
|---|---|---|---|
| | in school beginners of a German city (Frankfurt/Main)] | | |
| Schneider 2004 | Pathways to school achievement in very preterm and full term children | https://dx.doi.org/10.1007/BF03173217 | Wrong intervention |
| Schraeder 1987 | Preschool development of very low birthweight infants | https://dx.doi.org/10.1111/j.1547-5069.1987.tb00002.x | Wrong intervention |
| Schubiger 1999 | [Development of formerly preterm infants with a birth weight of below 1500 grams: concept and results of a follow-up program in up to school-aged children in central Switzerland] | | Wrong intervention |
| Sejer 2019 | Impact of gestational age on child intelligence, attention and executive function at age 5: a cohort study | https://dx.doi.org/10.1136/bmjopen-2019-028982 | Wrong intervention |
| Shah 2016 | Developmental Outcomes of Late Preterm Infants From Infancy to Kindergarten | https://dx.doi.org/10.1542/peds.2015-3496 | Wrong intervention |
| Shah 2016 | Gestational Age and Kindergarten School Readiness in a National Sample of Preterm Infants | https://dx.doi.org/10.1016/j.jpeds.2016.06.062 | Wrong intervention |
| Shah 2024 | Developmental trajectories of late preterm infants and predictors of academic performance | https://dx.doi.org/10.1038/s41390-023-02756-2 | Wrong intervention |
| Sheridan 1987 | Prematures enter school: a follow-up study | | Wrong intervention |
| Skranes 2017 | Executive function deficits in preterm subjects are a combination of social risk factors and brain maldevelopment | https://dx.doi.org/10.1111/apa.13955 | Wrong intervention |
| Smith-Longee 2024 | The early educational environment at five years of age in a European cohort of children born very preterm: challenges and opportunities for research | https://dx.doi.org/10.1186/s12887-024-04792-1 | Wrong study design |
| Snyder 2007 | Examining attention networks in preschool children born with very low birth weights | | Wrong intervention |
| Spiel 1990 | Children at biological risk in stressful situations: The influence of day care providers | | Wrong intervention |
| Squarza 2016 | Learning Disabilities in Extremely Low Birth Weight Children and Neurodevelopmental Profiles at Preschool Age | https://dx.doi.org/10.3389/fpsyg.2016.00998 | Wrong intervention |
| Stampoltzis | Developmental, familial and educational | https://dx.doi.org/10.1016/j.rasd.2012.05.004 | Wrong patient |

| | | | |
|---|---|---|---|
| 2012 | characteristics of a sample of children with Autism Spectrum Disorders in Greece | | population |
| Stephenson 2024 | Childcare use and the social-emotional and behavioural outcomes of late-preterm and early-term born children at age 5: An analysis of the All Our Families longitudinal cohort | https://dx.doi.org/10.17269/s41997-024-00908-3 | Wrong intervention |
| Sticker 1985 | Comparison of school performance between premature and full-term children up to the age of 13 years | | Wrong intervention |
| Sticker 1985 | Born too early-disadvantageous for school achievement? | | Wrong study design |
| Streiftau 2014 | Behavioural problems and learning impairments at age 7-10 after extreme prematurity | https://dx.doi.org/10.1026/0942-5403/a000149 | Wrong intervention |
| Sullivan 2003 | Perinatal morbidity, mild motor delay, and later school outcomes | | Wrong intervention |
| Svobodova 1967 | [School progress as a criterion for the mental development of immature children] | | Wrong language |
| Takeuchi 2016 | Reading difficulty in school-aged very low birth weight infants in Japan | https://dx.doi.org/10.1016/j.braindev.2016.04.013 | Wrong intervention |
| Tallqvist 1959 | Morbidity of prematures. An investigation based on questionnaires sent to the parents of children attending elementary school | | Wrong patient population |
| Tamiasso Vieira 2017 | Functional capacity, independence and home affordances of premature children attending daycare centers | 10.1590/1980-5918.030.001.AO09 | Wrong intervention |
| Taskila 2022 | Antenatal and neonatal risk factors in very preterm children were associated with language difficulties at 9 years of age | https://dx.doi.org/10.1111/apa.16501 | Wrong intervention |
| Tatsuoka 2016 | Effects of Extreme Prematurity on Numerical Skills and Executive Function in Kindergarten Children: An Application of Partially Ordered Classification Modeling | https://dx.doi.org/10.1016/j.lindif.2016.05.002 | Wrong intervention |
| Taylor 2019 | Associations of attention deficit hyperactivity disorder (ADHD) at school entry with early academic progress in children born prematurely and full-term controls | https://dx.doi.org/10.1016/j.lindif.2018.10.008 | Wrong intervention |

| | | | |
|---|---|---|---|
| Taylor 2024 | School Readiness Predictors of Early Academic Achievement in Children Born Very Preterm | https://dx.doi.org/10.1097/DBP.0000000000001275 | Wrong intervention |
| Taylor 1995 | Achievement in children with birth weights less than 750 grams with normal cognitive abilities: evidence for specific learning disabilities | https://dx.doi.org/10.1093/jpepsy/20.6.703 | Wrong intervention |
| Taylor 2011 | Learning problems in kindergarten students with extremely preterm birth | https://dx.doi.org/10.1001/archpediatrics.2011.137 | Wrong intervention |
| Taylor 2018 | Effects of extreme prematurity and kindergarten neuropsychological skills on early academic progress | https://dx.doi.org/10.1037/neu0000434 | Wrong study design |
| Taylor 2019 | Resilience in Extremely Preterm/Extremely Low Birth Weight Kindergarten Children | https://dx.doi.org/10.1017/S1355617719000080 | Wrong intervention |
| Taylor 2022 | School Readiness in 4-Year-Old Very Preterm Children | https://dx.doi.org/10.3390/children9030323 | Wrong intervention |
| Taylor 2016 | A simple screen performed at school entry can predict academic under-achievement at age seven in children born very preterm | https://dx.doi.org/10.1111/jpc.13186 | Wrong intervention |
| Tough 2008 | Maternal mental health predicts risk of developmental problems at 3 years of age: follow up of a community based trial | https://dx.doi.org/10.1186/1471-2393-8-16 | Wrong patient population |
| Towers 2018 | What are the outcomes for children born preterm and how can interventions meet their needs? | https://dx.doi.org/10.1080/02667363.2018.1426557 | Wrong study design |
| Townley Flores 2021 | Short-Term and Long-Term Educational Outcomes of Infants Born Moderately and Late Preterm | https://dx.doi.org/10.1016/j.jpeds.2020.12.070 | Wrong intervention |
| van Baar 2006 | Developmental Course of Very Preterm Children in Relation to School Outcome | https://dx.doi.org/10.1007/s10882-006-9016-6 | Wrong intervention |
| van Baar 2009 | Functioning at school age of moderately preterm children born at 32 to 36 weeks' gestational age | https://dx.doi.org/10.1542/peds.2008-2315 | Wrong intervention |
| van Beek 2021 | The Need for Special Education Among ELBW and SGA Preterm Children: A Cohort Study | https://dx.doi.org/10.3389/fped.2021.719048 | Wrong intervention |
| van de Bor 2004 | School performance in adolescents with and without periventricular-intraventricular hemorrhage in the neonatal period | https://dx.doi.org/10.1053/j.semperi.2004.08.007 | Wrong intervention |
| van de Weijer-Bergsma 2008 | Attention development in infants and preschool children born preterm: a review | https://dx.doi.org/10.1016/j.infbeh.2007.12.003 | Wrong study design |

| | | | |
|---|---|---|---|
| van Kessel-Feddema 2007 | Concordance between school outcomes and developmental follow-up results of very preterm and/or low birth weight children at the age of 5 years | https://dx.doi.org/10.1007/s00431-006-0309-7 | Wrong intervention |
| van Veen 2018 | Very preterm born children at early school age: Healthcare therapies and educational provisions | https://dx.doi.org/10.1016/j.earlhumdev.2017.12.010 | Wrong intervention |
| Vasconcellos 2022 | Prematurity, Family Environment Linked to Lower Rate of School Readiness | | Wrong intervention |
| Vekerdy 1991 | [Follow up of extremely low birth weight infants at the age of 8-11 years (late prognosis of perinatal factors)] | | Wrong language |
| Vekerdy-Lakatos 1989 | Infants weighing 1,000 g or less at birth. Outcome at 8-11 years of age | https://dx.doi.org/10.1111/j.1651-2227.1989.tb11284.x | Wrong intervention |
| Verkerk 2016 | Attention in 3-Year-Old Children with VLBW and Relationships with Early School Outcomes | https://dx.doi.org/10.3109/01942638.2015.1012319 | Wrong intervention |
| Verkerk 2014 | The relationship between multiple developmental difficulties in very low birth weight children at 31/2 years of age and the need for learning support at 5 years of age | https://dx.doi.org/10.1016/j.ridd.2013.10.007 | Wrong intervention |
| Verkerk 2013 | Assessing independency in daily activities in very preterm children at preschool age | https://dx.doi.org/10.1016/j.ridd.2013.03.032 | Wrong intervention |
| Vohr 1985 | Neurodevelopmental and school performance of very low-birth-weight infants: a seven-year longitudinal study | | Wrong intervention |
| Volgina 2001 | [The course of physical development of children born premature] | | Wrong language |
| Wakeley 2003 | Early mathematical development in very low-birthweight children | | Wrong study design |
| Walczak-Kozlowska 2022 | Heterogeneity of the attentional system's efficiency among very prematurely born pre-schoolers | https://dx.doi.org/10.1080/09297049.2021.1961702 | Wrong intervention |
| Wasson 2000 | Predicting language and behavioural outcomes for high-risk children at eight years of age | | Wrong intervention |
| Weindrich 2003 | Late sequelae of low birthweight: mediators of poor school performance at 11 years | 10.1017/s0012162203000860 | Wrong intervention |
| Wenstrom 2008 | [Commentary on] School outcomes of late | | Wrong intervention |

| | | | |
|---|---|---|---|
| | preterm infants: special needs and challenges for infants born at 32- to 36-week gestation | | |
| Wiener 1968 | Scholastic achievement at age 12-13 of prematurely born infants | https://dx.doi.org/10.1177/002246696800200302 | Wrong intervention |
| Winchester 2009 | Academic, social, and behavioral outcomes at age 12 of infants born preterm | https://dx.doi.org/10.1177/0193945909339321 | Wrong intervention |
| Wocadlo 2007 | Phonology, rapid naming and academic achievement in very preterm children at eight years of age | https://dx.doi.org/10.1016/j.earlhumdev.2006.08.001 | Wrong intervention |
| Wolke 2008 | Specific language difficulties and school achievement in children born at 25 weeks of gestation or less | https://dx.doi.org/10.1016/j.jpeds.2007.06.043 | Wrong intervention |
| Alenius 2006 | School grades and educational attainments of adolescents and young adults born preterm | https://dx.doi.org/10.1038/s41598-022-27295-5 | Wrong intervention |
| Andrews 2006 | Prediction of special education placement from birth certificate data | | Wrong patient population |

**Supplemental-Figure S 1.** PRISMA Flow Chart[4]

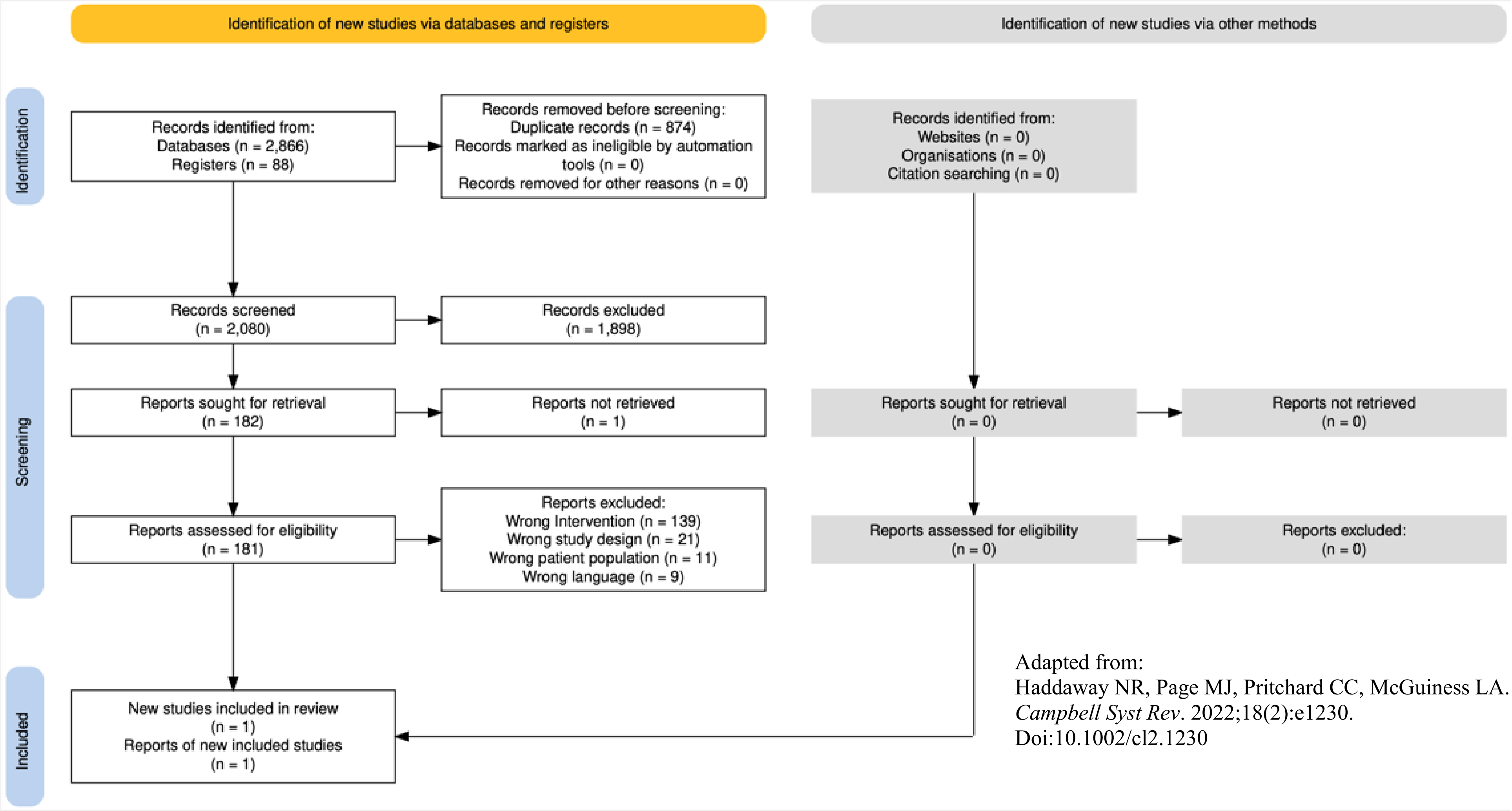


Adapted from:
Haddaway NR, Page MJ, Pritchard CC, McGuiness LA. *Campbell Syst Rev*. 2022;18(2):e1230. Doi:10.1002/cl2.1230

**Supplemental-Table S 4.** Risk of bias ratings

| **Study ID** | **Intervention** | **Comparator** | **Outcome** | **D1[a,b]** | **D2[c]** | **D3[d]** | **D4[e]** | **D5[f]** | **D6[g]** | **D7[h]** | **Overall** |
|---|---|---|---|---|---|---|---|---|---|---|---|
| Jaekel, 2015 | DSE | ASE | reading (teacher ratings) | 🟡 | 🟢 | 🟢 | 🟢 | 🟢 | 🔴 | 🔴 | 🔴 |
| Jaekel, 2015 | DSE | ASE | writing (teacher ratings) | 🟡 | 🟢 | 🟢 | 🟢 | 🟢 | 🔴 | 🔴 | 🔴 |
| Jaekel, 2015 | DSE | ASE | mathematics (teacher ratings) | 🟡 | 🟢 | 🟢 | 🟢 | 🟢 | 🔴 | 🔴 | 🔴 |
| Jaekel, 2015 | DSE | ASE | attention (teacher ratings) | 🟡 | 🟢 | 🟢 | 🟢 | 🟢 | 🔴 | 🔴 | 🔴 |
| Jaekel, 2015 | DSE | ASE | reading (Zürich- and Pseudowort Reading test) | 🟡 | 🟢 | 🟢 | 🟢 | 🟢 | 🟢 | 🟢 | 🟢 |
| Jaekel, 2015 | DSE | ASE | writing (diagnostic spelling test) | 🟡 | 🟢 | 🟢 | 🟢 | 🟢 | 🟢 | 🟢 | 🟢 |
| Jaekel, 2015 | DSE | ASE | mathematics (standardized mathematical test) | 🟡 | 🟢 | 🟢 | 🟢 | 🟢 | 🟢 | 🟢 | 🟢 |
| Jaekel, 2015 | DSE | ASE | attention (subscale of the Child Behavior and Rating Scale) | 🟡 | 🟢 | 🟢 | 🟢 | 🟢 | 🟢 | 🟢 | 🟢 |

🟢 Low Risk of Bias
🟡 Moderate Risk of Bias
🔴 Serious Risk of Bias
⚫ Critical Risk of Bias

[a]Bias due to confounding
[b]as not all confounders can usually be ruled out, this domain is typically rated as moderate risk, which may still allow for an overall low risk judgment.
[c]Bias in classification of interventions
[d]Bias in selection of participants into the study (or into the analysis)
[e]Bias due to deviations from intended interventions
[f]Bias due to missing data
[g]Bias in measurement of the outcome
[h]Bias in selection of the reported result
DSE – delayed school entry
ASE – age-appropriate school entry

**Supplemental-Table S 5.** Effects of interventions

| | | ASE | DSE | MD | B (95% CI) |
|---|---|---|---|---|---|
| **Reading** | Mean | 99.9 | 85.6 | 14.3 | -0.62 (-1.01 to -0.24) |
| **Writing** | | 100.5 | 85.8 | 14.7 | -0.98 (-1.17 to -0.78) |
| **Mathematics** | | 97.5 | 88.3 | 9.2 | -0.30 (-0.65 to 0.04) |
| **Attention** | | 100.1 | 83.8 | 16.3 | -0.74 (-1.05 to -0.43) |

Abbreviations:
ASE: age-appropriate school entry, B: regression coefficient, CI: confidence interval, DSE: delayed school entry, MD: mean difference

**Appendix 4. Drop-out analysis and analysis based on chronological age**

**Supplemental-Table S 6.** Drop-out-Analysis

| | | **included in GNN** | **Follow-up age 5 years** | **Follow-up age 10 years** |
|---|---|---|---|---|
| | | 13848 | 3403 (24.6) | 1869 (13.5) |
| **Gestational age** (weeks)* [b] | Mean (sd) | 28.0 (2.3) | 27.8 (2.2) | 27.8 (2.2) |
| **Birth weight** (grams)* [b] | | 1015 (301) | 983 (289) | 993 (287) |
| **Male** [a] | n (%) | 7208 (52.1) | 1773 (52.1) | 963 (51.5) |
| **BPD*** [a] | | 2616 (18.9) | 687 (20.2) | 376 (20.1) |
| **IVH** [a] | | 2678 (19.4) | 630 (18.5) | 358 (19.2) |
| grade I | | 1067 (7.7) | 244 (7.2) | 141 (7.5) |
| grade II | | 695 (5.0) | 174 (5.1) | 99 (5.3) |
| grade III | | 460 (3.3) | 112 (3.3) | 54 (2.9) |
| grade IV | | 455 (3.3) | 99 (2.9) | 63 (3.4) |
| **Non-german heritage*** [a] | | 3763 (27.2) | 660 (19.4) | 229 (12.3) |

* $p<0.008$ (corrected for multiple testing by Bonferroni correction)
[a] tested by Chi-square test.
[b] tested by Mann-Whitney U test
Abbreviations:
GNN: German Neontal Network, BPD: Bronchopulmonary Dysplasia, IVH: Intraventricular Hemorrhage, mean: mean value, sd: standard deviation, n number

**Supplemental-Table S 7.** Baseline characteristics (chronological age)

| | | **ASE** | **DSE** | **p-Value** |
|---|---|---|---|---|
| | | 1420 (87.2) | 200 (12.3) | |
| **GA** (weeks) | Mean (sd) | 28.1 (2.0) | 27.1 (2.0) | <0.001 [b] |
| **Birth weight** (grams) | | 1034 (275 | 895 (283) | <0.001 [b] |
| **Male** | n (%) | 690 (48.6) | 121 (61) | 0.002 [a] |
| **BPD** | | 200 (14.1) | 62 (31) | <0.001 [a] |
| **IVH** | | 229 (16.1) | 48 (24) | 0.006 [a] |
| grade I | | 111 (7.8) | 16 (8) | 0.007 [a] |
| grade II | | 64 (4.5) | 15 (8) | |
| grade III | | 27 (1.9) | 7 (4) | |
| grade IV | | 27 (1.9) | 10 (5) | |
| **Cerebral palsy** | | 32 (2.4) | 6 (3) | 0.51 [a] |
| **Mother high school** | | 756 (55.7) | 93 (50) | 0.12 [a] |
| **IQ** at age 5 | Mean (sd) | 102 (11) | 95 (12) | <0.001 [b] |
| **SDQ** at age 5 | | 8.5 (5.2) | 10.7 (5.8) | <0.001 [b] |
| **MABC** at age 5 | | 9.4 (3.1) | 7.5 (2.8) | <0.001 [b] |
| **Club sport** at age 5 | n (%) | 818 (57.6) | 91 (46) | <0.001 [a] |

[a] tested by Chi-square test.
[b] tested by Mann-Whitney U test.
Abbreviations
ASE: age-appropriate school entry, DSE: delayed school entry, BPD: Bronchopulmonary Dysplasia, IQ: Intelligence Quotient, IVH: Intraventricular Hemorrhage, MABC: Movement Assessment Battery for Children- II, SDQ: Strengths and Difficulties Questionnaire, mean: mean value, sd: standard deviation, n number

**Supplemental-Table S 8.** Outcomes of appropriate versus delayed school entry with regard to F-words at 10 years of age (chronological age)

| | | ASE | DSE | p-Value |
|---|---|---|---|---|
| **Function** | | | | |
| Special educational needs | n (%) | 131 (9.2) | 48 (24) | <0.001 [a] |
| Executive functions (BRIEF, *PR*) | Mean (sd) | 52 (11) | 56 (12) | <0.001 [b] |
| Attention (SDQ, *SR*) | | 3.6 (1.2) | 3.9 (1.2) | <0.001 [b] |
| Internalizing problems (SDQ, *SR*) | | 4.5 (3.2) | 5.3 (3.8) | 0.02 [b] |
| Cognitive processing (KOPKIJ, *PR*) | | | | |
| Speech | | 1.2 (0.2) | 1.2 (0.3) | <0.001 [b] |
| Memory | | 1.4 (0.4) | 1.5 (0.5) | <0.001 [b] |
| Visual-spatial | | 1.4 (0.5) | 1.6 (0.6) | <0.001 [b] |
| School | | 1.44 (0.5) | 1.7 (0.6) | <0.001 [b] |
| **Friendship (KIDSCREEN-52. *SR*)** | | | | |
| Peer | | 50.9 (10.0) | 49.7 (10.6) | 0.04 [b] |
| Bullying | | 49.4 (10.2) | 48.9 (10.8) | 0.713 [b] |
| **Family (KIDSCREEN-52. *SR*)** | | | | |
| Parents | | 54.4 (7.5) | 53.2 (8.0) | 0.11 [b] |
| **Fun (KIDSCREEN-52, *PR*)** | | | | |
| Autonomy | | 53.2 (7.9) | 52.5 (8.2) | 0.31 [b] |
| **Fitness** | | | | |
| Club sports | n (%) | 1086 (84.4) | 142 (79) | 0.04 [a] |
| **Participation (CASP, *PR*)** | Mean (sd) | 96.0 (8.2) | 92.1 (12.6) | <0.001 [b] |
| **Quality of life (KIDSCREEN-52, *SR*)** | | 54.0 (6.1) | 53.2 (6.7) | 0.18 [b] |

[a] tested by Chi-square test.
[b] tested by Mann-Whitney U test.
Abbreviations:
ASE: age-appropriate school entry, DSE: delayed school entry, mean: mean value, sd: standard deviation, n: number, CASP: Child and Adolescent Scale of Participation, SEN: special educational needs, BRIEF: Behavior Rating Inventory of Executive Function, SDQ: Strength and Difficulties Questionnaire, KOPKIJ: Questionnaire for the Assessment of Cognitive Problems in Children and Adolescents, PR: parental report, SR: self report

**Supplemental-Table S 9.** Effects of Delayed School entry (DSE) based on chronological age

| | **B** | **95%-CI** | | **p-Value** |
|---|---|---|---|---|
| **Function** | | | | |
| Executive functions (BRIEF, *PR*) | -0.023 | -1.824 | 1.778 | 0.98 |
| Attention (SDQ, *SR*) | -0.066 | -0.294 | 0.162 | 0.57 |
| internalizing problems (SDQ, *SR*) | -0.007 | -0.654 | 0.639 | 0.98 |
| <u>Cognitive Processing (KOPKIJ, *PR*)</u> | | | | |
| Speech | 0.033 | -0.012 | 0.078 | 0.16 |
| Memory | 0.017 | -0.059 | 0.093 | 0.66 |
| visual-spatial | 0.066 | -0.023 | 0.154 | 0.15 |
| School | 0.104 | 0.004 | 0.203 | 0.04 |
| **Friendship (KIDSCREEN-52, *SR*)** | | | | |
| Peers | -0.301 | -2.327 | 1.724 | 0.77 |
| Bullying | 1.791 | -0.349 | 3.931 | 0.10 |
| **Family (KIDSCREEN-52, *SR*)** | | | | |
| Parents | -0.5 | -2.116 | 1.117 | 0.54 |
| **Fun (KIDSCREEN-52, *PR*)** | | | | |
| Autonomy | 0.734 | -0.858 | 2.326 | 0.37 |
| **Participation (CASP, *PR*)** | -0.646 | -1.997 | 0.704 | 0.35 |
| **Quality of life (KIDSCREEN-52, *SR*)** | 0.485 | -0.770 | 1.740 | 0.45 |
| | **OR** | **95%-CI** | | **p-Value** |
| **Function** | | | | |
| SEN | 1.018 | 0.547 | 1.896 | 0.96 |
| **Fitness** | | | | |
| Club Sports | 1.550 | 0.801 | 3.001 | 0.19 |

<u>Abbreviations:</u>
DSE: delayed school entry, CASP: Child and Adolescent Scale of Participation, SEN: special educational needs, BRIEF: Behavior Rating Inventory of Executive Function, SDQ: Strength and Difficulties Questionnaire, KOPKIJ: Questionnaire for the Assessment of Cognitive Problems in Children and Adolescents, B: regression coefficient, CI: confidence interval, OR: Odds ratio, PR: parental report, SR: self report
<u>Adjusted linear and logistic regression for:</u> gestational age, birth weight, gender, bronchopulmonary dysplasia, intraventricular hemorrhage, mother high school, cerebral palsy, at age of 5: IQ, Movement Assessment Battery for Children, Stength and difficulties Questionnaire, club sports

**Appendix 4. Total regression analysis (based on chronological age)**

**Supplemental-Table S 10.** Special educational needs

| | **B** | **S.E.** | **p-Value** | **Odds Ratio** | **95%-CI** |
|---|---|---|---|---|---|
| **Delayed school entry** | 0.018 | 0.317 | 0.96 | 1.018 | 0.547-1.896 |
| **Gestational age** | 0.065 | 0.090 | 0.46 | 1.068 | 0.896-1.273 |
| **Birth weight** | 0.000 | 0.001 | 0.61 | 1.000 | 0.998-1.001 |
| **Male sex** | 0.085 | 0.266 | 0.75 | 1.088 | 0.646-1.833 |
| **BPD** | 0.014 | 0.321 | 0.96 | 1.014 | 0.540-1.904 |
| **IVH** | 0.164 | 0.327 | 0.62 | 1.178 | 0.620-2.236 |
| **Mother high school** | 0.054 | 0.264 | 0.84 | 1.056 | 0.630-1.769 |
| **Cerebral palsy** | -0.217 | 1.364 | 0.87 | 0.805 | 0.056-11.659 |
| **IQ** at age 5 years | -0.075 | 0.013 | <0.001 | 0.927 | 0.904-0.951 |
| **MABC** at age 5 years | -0.128 | 0.049 | 0.009 | 0.880 | 0.800-0.968 |
| **Clubsports** at age 5 years | -0.666 | 0.256 | 0.009 | 0.514 | 0.311-0.848 |
| **SDQ** at age 5 years | 0.134 | 0.023 | <0.001 | 1.143 | 1.094-1.194 |

Abbreviation:
BPD: Bronchopulmonary Dysplasia, IQ: Intelligence Quotient, IVH: Intraventricular Hemorrhage, MABC: Movement Assessment Battery for Children- II, SDQ: Strengths and Difficulties Questionnaire, S.E.: standard error, B: Regressioncoefficient, CI: Confidence interval

**Supplemental-Table S 11** Clubsports

| | B | S.E. | p-Value | Odds Ratio | 95%-CI |
|---|---|---|---|---|---|
| **Delayed school entry** | 0.438 | 0.337 | 0.20 | 1.550 | 0.801-3.001 |
| **Gestational age** | -0.048 | 0.080 | 0.55 | 0.953 | 0.815-1.115 |
| **Birth weight** | 0.000 | 0.001 | 0.66 | 1.000 | 0.999-1.001 |
| **Male sex** | -0.334 | 0.223 | 0.14 | 0.716 | 0.463-1.109 |
| **BPD** | 0.659 | 0.329 | 0.05 | 1.934 | 1.014-3.687 |
| **IVH** | 0.056 | 0.314 | 0.858 | 1.058 | 0.571-1.958 |
| **Mother high school** | 0.492 | 0.220 | 0.03 | 1.636 | 1.064-2.516 |
| **Cerebral palsy** | -0.772 | 1.278 | 0.55 | 0.462 | 0.038-5.658 |
| **IQ** at age 5 years | 0.059 | 0.010 | <0.001 | 1.060 | 1.039-1.082 |
| **MABC** at age 5 years | 0.060 | 0.040 | 0.13 | 1.062 | 0.983-1.149 |
| **Clubsports** at age 5 years | 1.698 | 0.233 | <0.001 | 5.462 | 3.458-8.627 |
| **SDQ** at age 5 years | 0.013 | 0.021 | 0.56 | 1.013 | 0.971-1.056 |

Abbreviation:
BPD: Bronchopulmonary Dysplasia, IQ: Intelligence Quotient, IVH: Intraventricular Hemorrhage, MABC: Movement Assessment Battery for Children- II, SDQ: Strengths and Difficulties Questionnaire, S.E.: standard error, B: Regression coefficient, CI: Confidence interval

**Supplemental-Table S 12.** Participation (CASP, parental reported)

| | B | S.E. | p-Value | 95%-CI | |
|---|---|---|---|---|---|
| **Delayed school entry** | -0.646 | 0.688 | 0.35 | -1.997 | 0.704 |
| **Gestational age** | 0.022 | 0.164 | 0.90 | -0.300 | 0.343 |
| **Birth weight** | 0.001 | 0.001 | 0.54 | -0.002 | 0.003 |
| **Male sex** | 0.703 | 0.454 | 0.12 | -0.188 | 1.594 |
| **BPD** | 0.093 | 0.641 | 0.89 | -1.166 | 1.351 |
| **IVH** | 0.013 | 0.646 | 0.98 | -1.255 | 1.280 |
| **Mother high school** | -0.243 | 0.448 | 0.59 | -1.123 | 0.637 |
| **Cerebral palsy** | -7.413 | 3.803 | 0.05 | -14.877 | 0.050 |
| **IQ** at age 5 years | 0.082 | 0.021 | <0.001 | 0.041 | 0.123 |
| **MABC** at age 5 years | 0.347 | 0.080 | <0.001 | 0.191 | 0.504 |
| **Clubsports** at age 5 years | 1.006 | 0.448 | 0.03 | 0.127 | 1.885 |
| **SDQ** at age 5 years | -0.576 | 0.043 | <0.001 | -0.661 | -0.490 |

Abbreviation:
BPD: Bronchopulmonary Dysplasia, IQ: Intelligence Quotient, IVH: Intraventricular Hemorrhage, MABC: Movement Assessment Battery for Children- II, SDQ: Strengths and Difficulties Questionnaire, S.E.: standard error, B: Regression coefficient, CI: Confidence interval

**Supplemental-Table S 13.** Quality of life (KIDSCREEN-52, self reported)

| | B | S.E. | p-Value | 95%-CI | |
|---|---|---|---|---|---|
| **Delayed school entry** | 0.485 | 0.639 | 0.45 | -0.770 | 1.740 |
| **Gestational age** | -0.131 | 0.149 | 0.38 | -0.424 | 0.162 |
| **Birth weight** | 0.000 | 0.001 | 0.86 | -0.002 | 0.002 |
| **Male sex** | -0.590 | 0.418 | 0.16 | -1.411 | 0.231 |
| **BPD** | 0.225 | 0.593 | 0.70 | -0.938 | 1.389 |
| **IVH** | -0.867 | 0.584 | 0.14 | -2.013 | 0.280 |
| **Mother high school** | -0.019 | 0.413 | 0.96 | -0.830 | 0.793 |
| **Cerebral palsy** | 2.401 | 3.465 | 0.49 | -4.399 | 9.202 |
| **IQ** at age 5 years | 0.023 | 0.019 | 0.22 | -0.014 | 0.061 |
| **MABC** at age 5 years | 0.095 | 0.074 | 0.20 | -0.049 | 0.240 |
| **Clubsports** at age 5 years | 0.476 | 0.411 | 0.25 | -0.331 | 1.282 |
| **SDQ** at age 5 years | -0.340 | 0.040 | <0.001 | -0.419 | -0.261 |

Abbreviation:
BPD: Bronchopulmonary Dysplasia, IQ: Intelligence Quotient, IVH: Intraventricular Hemorrhage, MABC: Movement Assessment Battery for Children- II, SDQ: Strengths and Difficulties Questionnaire, S.E.: standard error, B: Regression coefficient, CI: Confidence interval

**Supplemental-Table S 14.** Bullying (KIDSCREEN-52, self reported

| | B | S.E. | p-Value | 95%-CI | |
|---|---|---|---|---|---|
| **Delayed school entry** | 1.791 | 1.090 | 0.10 | -0.349 | 3.931 |
| **Gestational age** | -0.499 | 0.254 | 0.05 | -0.999 | 0.000 |
| **Birth weight** | 0.003 | 0.002 | 0.15 | -0.001 | 0.006 |
| **Male sex** | -0.281 | 0.713 | 0.69 | -1.681 | 1.120 |
| **BPD** | 0.747 | 1.011 | 0.46 | -1.238 | 2.731 |
| **IVH** | -2.433 | 0.996 | 0.02 | -4.388 | -0.478 |
| **Mother high school** | 0.344 | 0.705 | 0.626 | -1.040 | 1.727 |
| **Cerebral palsy** | 5.154 | 5.909 | 0.383 | -6.443 | 16.750 |
| **IQ** at age 5 years | 0.076 | 0.033 | 0.019 | 0.013 | 0.140 |
| **MABC** at age 5 years | 0.119 | 0.125 | 0.343 | -0.127 | 0.365 |
| **Clubsports** at age 5 years | -0.412 | 0.701 | 0.557 | -1.788 | 0.964 |
| **SDQ** at age 5 years | -0.461 | 0.069 | <0.001 | -0.569 | -0.326 |

Abbreviation:
BPD: Bronchopulmonary Dysplasia, IQ: Intelligence Quotient, IVH: Intraventricular Hemorrhage, MABC: Movement Assessment Battery for Children- II, SDQ: Strengths and Difficulties Questionnaire, S.E.: standard error, B: Regression coefficient, CI: Confidence interval

**Supplemental-Table S 15.** Attention (SDQ, self reported)

| | B | S.E. | p-Value | 95%-CI | |
|---|---|---|---|---|---|
| **Delayed school entry** | -0.066 | 0.116 | 0.57 | -0.294 | 0.162 |
| **Gestational age** | 0.016 | 0.027 | 0.57 | -0.038 | 0.069 |
| **Birth weight** | 0.000 | 0.000 | 0.13 | -0.001 | 0.000 |
| **Male sex** | -0.164 | 0.076 | 0.03 | -0.314 | -0.015 |
| **BPD** | -0.139 | 0.108 | 0.20 | -0.351 | 0.073 |
| **IVH** | 0.035 | 0.106 | 0.74 | -0.174 | 0.243 |
| **Mother high school** | -0.141 | 0.075 | 0.062 | -0.288 | 0.007 |
| **Cerebral palsy** | 0.089 | 0.631 | 0.89 | -1.149 | 1.327 |
| **IQ** at age 5 years | -0.012 | 0.003 | 0.001 | -0.019 | -0.005 |
| **MABC** at age 5 years | -0.025 | 0.013 | 0.07 | -0.051 | 0.002 |
| **Clubsports** at age 5 years | -0.019 | 0.075 | 0.80 | -0.166 | 0.128 |
| **SDQ** at age 5 years | 0.081 | 0.007 | <0.001 | 0.067 | 0.096 |

Abbreviation:
BPD: Bronchopulmonary Dysplasia, IQ: Intelligence Quotient, IVH: Intraventricular Hemorrhage, MABC: Movement Assessment Battery for Children- II, SDQ: Strengths and Difficulties Questionnaire, S.E.: standard error, B: Regression coefficient, CI: Confidence interval

**Supplemental-Table S 16.** Internalizing problems (SDQ, self reported)

| | B | S.E. | p-Value | 95%-CI | |
|---|---|---|---|---|---|
| **Delayed school entry** | -0.007 | 0.329 | 0.98 | -0.654 | 0.639 |
| **Gestational age** | 0.125 | 0.077 | 0.10 | -0.025 | 0.276 |
| **Birth weight** | -0.001 | 0.001 | 0.17 | -0.002 | 0.000 |
| **Male sex** | 0.840 | 0.215 | <0.001 | 0.418 | 1.263 |
| **BPD** | 0.248 | 0.305 | 0.42 | -0.351 | 0.847 |
| **IVH** | 0.509 | 0.301 | 0.09 | -0.081 | 1.100 |
| **Mother high school** | -0.013 | 0.213 | 0.95 | -0.431 | 0.405 |
| **Cerebral palsy** | -0.402 | 1.784 | 0.822 | -3.905 | 3.100 |
| **IQ** at age 5 years | -0.007 | 0.010 | 0.50 | -0.026 | 0.013 |
| **MABC** at age 5 years | -0.062 | 0.038 | 0.11 | -0.136 | 0.013 |
| **Clubsports** at age 5 years | -0.063 | 0.212 | 0.77 | -0.478 | 0.353 |
| **SDQ** at age 5 years | 0.241 | 0.021 | <0.001 | 0.200 | 0.282 |

Abbreviation:
BPD: Bronchopulmonary Dysplasia, IQ: Intelligence Quotient, IVH: Intraventricular Hemorrhage, MABC: Movement Assessment Battery for Children- II, SDQ: Strengths and Difficulties Questionnaire, S.E.: standard error, B: Regression coefficient, CI: Confidence interval

**Supplemental-Table S 17.** School (KOPKIJ, parental reported)

| | B | S.E. | p-Value | 95%-CI | |
|---|---|---|---|---|---|
| **Delayed school entry** | 0.104 | 0.050 | 0.04 | 0.004 | 0.203 |
| **Gestational age** | 0.017 | 0.012 | 0.16 | -0.007 | 0.041 |
| **Birth weight** | 0.000 | 0.000 | 0.47 | 0.000 | 0.000 |
| **Male sex** | 0.005 | 0.033 | 0.89 | -0.061 | 0.070 |
| **BPD** | -0.021 | 0.047 | 0.67 | -0.114 | 0.073 |
| **IVH** | -0.009 | 0.047 | 0.84 | -0.102 | 0.083 |
| **Mother high school** | -0.051 | 0.033 | 0.12 | -0.116 | 0.014 |
| **Cerebral palsy** | -0.445 | 0.289 | 0.12 | -1.013 | 0.122 |
| **IQ** at age 5 years | -0.011 | 0.002 | <0.001 | -0.014 | -0.008 |
| **MABC** at age 5 years | -0.017 | 0.006 | 0.005 | -0.028 | -0.005 |
| **Clubsports** at age 5 years | -0.054 | 0.033 | 0.10 | -0.119 | 0.011 |
| **SDQ** at age 5 years | 0.028 | 0.003 | <0.001 | 0.022 | 0.035 |

Abbreviation:
BPD: Bronchopulmonary Dysplasia, IQ: Intelligence Quotient, IVH: Intraventricular Hemorrhage, MABC: Movement Assessment Battery for Children- II, SDQ: Strengths and Difficulties Questionnaire, S.E.: standard error, B: Regression coefficient, CI: Confidence interval

**Supplemental-Table S 18.** Executive function (BRIEF, parental reported)

| | B | S.E. | p-Value | 95%-CI | |
|---|---|---|---|---|---|
| **Delayed school entry** | -0.023 | 0.918 | 0.98 | -1.824 | 1.778 |
| **Gestational age** | 0.084 | 0.221 | 0.70 | -0.350 | 0.517 |
| **Birth weight** | -0.003 | 0.002 | 0.08 | -0.006 | 0.000 |
| **Male sex** | 1.017 | 0.611 | 0.10 | -0.181 | 2.216 |
| **BPD** | -2.366 | 0.863 | 0.006 | -4.060 | -0.673 |
| **IVH** | -0.692 | 0.859 | 0.42 | -2.378 | 0.995 |
| **Mother high school** | 0.387 | 0.604 | 0.52 | -0.799 | 1.573 |
| **Cerebral palsy** | -10.124 | 5.284 | 0.06 | -20.495 | 0.246 |
| **IQ** at age 5 years | -0.069 | 0.028 | 0.014 | -0.124 | -0.014 |
| **MABC** at age 5 years | -0.392 | 0.107 | <0.001 | -0.602 | -0.182 |
| **Clubsports** at age 5 years | -1.390 | 0.605 | 0.02 | -2.576 | -0.203 |
| **SDQ** at age 5 years | 1.069 | 0.059 | <0.001 | 0.954 | 1.183 |

Abbreviation:
BPD: Bronchopulmonary Dysplasia, IQ: Intelligence Quotient, IVH: Intraventricular Hemorrhage, MABC: Movement Assessment Battery for Children- II, SDQ: Strengths and Difficulties Questionnaire, S.E.: standard error, B: Regression coefficient, CI: Confidence interval

**Supplemental-Table S 19.** Speech (KOPKIJ, parental reported)

| | B | S.E. | p-Value | 95%-CI | |
|---|---|---|---|---|---|
| **Delayed school entry** | 0.033 | 0.023 | 0.16 | -0.012 | 0.078 |
| **Gestational age** | 0.002 | 0.006 | 0.73 | -0.009 | 0.013 |
| **Birth weight** | 0.000 | 0.000 | 0.56 | 0.000 | 0.000 |
| **Male sex** | 0.003 | 0.015 | 0.87 | -0.027 | 0.033 |
| **BPD** | 0.004 | 0.022 | 0.84 | -0.038 | 0.047 |
| **IVH** | 0.005 | 0.021 | 0.81 | -0.037 | 0.047 |
| **Mother high school** | -0.027 | 0.015 | 0.07 | -0.057 | 0.002 |
| **Cerebral palsy** | -0.268 | 0.132 | 0.04 | -0.527 | -0.009 |
| **IQ** at age 5 years | -0.004 | 0.001 | <0.001 | -0.005 | -0.002 |
| **MABC** at age 5 years | -0.005 | 0.003 | 0.06 | -0.010 | 0.000 |
| **Clubsports** at age 5 years | -0.013 | 0.015 | 0.37 | -0.043 | 0.016 |
| **SDQ** at age 5 years | 0.011 | 0.001 | <0.001 | 0.008 | 0.014 |

Abbreviation:
BPD: Bronchopulmonary Dysplasia, IQ: Intelligence Quotient, IVH: Intraventricular Hemorrhage, MABC: Movement Assessment Battery for Children- II, SDQ: Strengths and Difficulties Questionnaire, S.E.: standard error, B: Regression coefficient, CI: Confidence interval

**Supplemental-Table S 20.** Memory (KOPKIJ, parental reported)

| | B | S.E. | p-Value | 95%-CI | |
|---|---|---|---|---|---|
| **Delayed school entry** | 0.017 | 0.039 | 0.66 | -0.059 | 0.093 |
| **Gestational age** | 0.013 | 0.009 | 0.15 | -0.005 | 0.032 |
| **Birth weight** | 0.000 | 0.000 | 0.22 | 0.000 | 0.000 |
| **Male sex** | -0.026 | 0.026 | 0.32 | -0.077 | 0.025 |
| **BPD** | -0.035 | 0.036 | 0.34 | -0.107 | 0.036 |
| **IVH** | -0.021 | 0.036 | 0.56 | -0.093 | 0.050 |
| **Mother high school** | -0.037 | 0.026 | 0.15 | -0.087 | 0.013 |
| **Cerebral palsy** | -0.327 | 0.223 | 0.14 | -0.765 | 0.110 |
| **IQ** at age 5 years | -0.006 | 0.001 | <0.001 | -0.008 | -0.003 |
| **MABC** at age 5 years | -0.007 | 0.005 | 0.15 | -0.015 | 0.002 |
| **Clubsports** at age 5 years | -0.073 | 0.026 | 0.005 | -0.123 | -0.022 |
| **SDQ** at age 5 years | 0.034 | 0.002 | <0.001 | 0.029 | 0.038 |

Abbreviation:
BPD: Bronchopulmonary Dysplasia, IQ: Intelligence Quotient, IVH: Intraventricular Hemorrhage, MABC: Movement Assessment Battery for Children- II, SDQ: Strengths and Difficulties Questionnaire, S.E.: standard error, B: Regression coefficient, CI: Confidence interval

**Supplemental-Table S 21.** Visual-spatial (KOPKIJ, parental reported)

| | B | S.E. | p-Value | 95%-CI | |
|---|---|---|---|---|---|
| **Delayed school entry** | 0.066 | 0.045 | 0.15 | -0.023 | 0.154 |
| **Gestational age** | -0.014 | 0.011 | 0.19 | -0.036 | 0.007 |
| **Birth weight** | 0.000 | 0.000 | 0.61 | 0.000 | 0.000 |
| **Male sex** | -0.186 | 0.030 | <0.001 | -0.245 | -0.127 |
| **BPD** | 0.052 | 0.042 | 0.22 | -0.031 | 0.135 |
| **IVH** | 0.009 | 0.042 | 0.82 | -0.073 | 0.092 |
| **Mother high school** | 0.018 | 0.030 | 0.54 | -0.040 | 0.076 |
| **Cerebral palsy** | 0.909 | 0.259 | <0.001 | 0.401 | 1.417 |
| **IQ** at age 5 years | -0.007 | 0.001 | <0.001 | -0.010 | -0.004 |
| **MABC** at age 5 years | -0.016 | 0.005 | 0.002 | -0.027 | -0.006 |
| **Clubsports** at age 5 years | 0.031 | 0.030 | 0.30 | -0.028 | 0.089 |
| **SDQ** at age 5 years | 0.016 | 0.003 | <0.001 | 0.010 | 0.022 |

Abbreviation:
BPD: Bronchopulmonary Dysplasia, IQ: Intelligence Quotient, IVH: Intraventricular Hemorrhage, MABC: Movement Assessment Battery for Children- II, SDQ: Strengths and Difficulties Questionnaire, S.E.: standard error, B: Regression coefficient, CI: Confidence interval

**Supplemental-Table S 22**. Parents (KIDSCREEN-52, self reported)

| | B | S.E. | p-Value | 95%-CI | |
|---|---|---|---|---|---|
| **Delayed school entry** | -0.500 | 0.824 | 0.54 | -2.116 | 1.117 |
| **Gestational age** | -0.176 | 0.193 | 0.36 | -0.554 | 0.203 |
| **Birth weight** | 0.000 | 0.001 | 0.74 | -0.002 | 0.003 |
| **Male sex** | -0.064 | 0.540 | 0.91 | -1.125 | 0.997 |
| **BPD** | 0.318 | 0.765 | 0.68 | -1.184 | 1.819 |
| **IVH** | -0.202 | 0.755 | 0.79 | -1.683 | 1.280 |
| **Mother high school** | -0.398 | 0.534 | 0.46 | -1.446 | 0.650 |
| **Cerebral palsy** | 1.316 | 4.478 | 1.77 | -7.437 | 10.105 |
| **IQ** at age 5 years | 0.014 | 0.025 | 0.58 | -0.035 | 0.062 |
| **MABC** at age 5 years | 0.047 | 0.095 | 0.63 | -0.140 | 0.233 |
| **Clubsports** at age 5 years | 0.984 | 0.531 | 0.06 | -0.058 | 2.025 |
| **SDQ** at age 5 years | -0.248 | 0.052 | <0.001 | -0.350 | -0.146 |

Abbreviation:
BPD: Bronchopulmonary Dysplasia, IQ: Intelligence Quotient, IVH: Intraventricular Hemorrhage, MABC: Movement Assessment Battery for Children- II, SDQ: Strengths and Difficulties Questionnaire, S.E.: standard error, B: Regression coefficient, CI: Confidence interval

**Supplemental-Table S 23.** Peers (KIDSCREEN-52, self reported)

| | B | S.E. | p-Value | 95%-CI | |
|---|---|---|---|---|---|
| **Delayed school entry** | -0.301 | 1.032 | 0.77 | -2.327 | 1.724 |
| **Gestational age** | -0.235 | 0.241 | 0.33 | -0.707 | 0.238 |
| **Birth weight** | 0.000 | 0.002 | 0.86 | -0.003 | 0.004 |
| **Male sex** | -0.208 | 0.675 | 0.76 | -1.534 | 1.118 |
| **BPD** | 0.476 | 0.957 | 0.62 | -1.403 | 2.355 |
| **IVH** | -0.598 | 0.943 | 0.53 | -2.450 | 1.253 |
| **Mother high school** | -0.192 | 0.667 | 0.77 | -1.502 | 1.118 |
| **Cerebral palsy** | -1.516 | 5.594 | 0.79 | -12.495 | 9.464 |
| **IQ** at age 5 years | 0.038 | 0.031 | 0.22 | -0.023 | 0.098 |
| **MABC** at age 5 years | 0.225 | 0.119 | 0.06 | -0.008 | 0.458 |
| **Clubsports** at age 5 years | 1.076 | 0.664 | 0.11 | -0.227 | 2.378 |
| **SDQ** at age 5 years | -0.429 | 0.065 | <0.001 | -0.556 | -0.301 |

Abbreviation:
BPD: Bronchopulmonary Dysplasia, IQ: Intelligence Quotient, IVH: Intraventricular Hemorrhage, MABC: Movement Assessment Battery for Children- II, SDQ: Strengths and Difficulties Questionnaire, S.E.: standard error, B: Regression coefficient, CI: Confidence interval

**Supplemental-Table S 24.** Autonomy (KIDSCREEN-52, parental reported)

| | B | S.E. | p-Value | 95%-CI | |
|---|---|---|---|---|---|
| **Delayed school entry** | 0.734 | 0.811 | 0.37 | -0.858 | 2.326 |
| **Gestational age** | -0.007 | 0.193 | 0.97 | -0.386 | 0.372 |
| **Birth weight** | -0.001 | 0.001 | 0.38 | -0.004 | 0.002 |
| **Male sex** | -0.878 | 0.535 | 0.10 | -1.927 | 0.172 |
| **BPD** | -0.086 | 0.756 | 0.91 | -1.569 | 1.397 |
| **IVH** | -0.683 | 0.761 | 0.37 | -2.177 | 0.810 |
| **Mother high school** | -0.667 | 0.528 | 0.21 | -1.704 | 0.369 |
| **Cerebral palsy** | -8.576 | 4.485 | 0.06 | -17.378 | 0.226 |
| **IQ** at age 5 years | -0.044 | 0.025 | 0.08 | -0.092 | 0.004 |
| **MABC** at age 5 years | 0.218 | 0.094 | 0.02 | 0.033 | 0.403 |
| **Clubsports** at age 5 years | -0.591 | 0.527 | 0.26 | -1.626 | 0.444 |
| **SDQ** at age 5 years | -0.324 | 0.051 | <0.001 | -0.425 | -0.224 |

Abbreviation:
BPD: Bronchopulmonary Dysplasia, IQ: Intelligence Quotient, IVH: Intraventricular Hemorrhage, MABC: Movement Assessment Battery for Children- II, SDQ: Strengths and Difficulties Questionnaire, S.E.: standard error, B: Regression coefficient, CI: Confidence interval

**Appendix 5. Total regression analysis (based on corrected age)**

**Supplemental-Table S 25.** Special educational needs.

| | **B** | **S.E.** | **p-Value** | **Odds Ratio** | **95%-CI** |
|---|---|---|---|---|---|
| **Delayed school entry** | 0.180 | 0.403 | 0.66 | 1.197 | 0.544-2.637 |
| **Gestational age** | 0.066 | 0.090 | 0.46 | 1.068 | 0.896-1.273 |
| **Birth weight** | 0.000 | 0.001 | 0.62 | 1.000 | 0.998-1.001 |
| **Male sex** | 0.096 | 0.265 | 0.72 | 1.100 | 0.654-1.851 |
| **BPD** | 0.017 | 0.321 | 0.96 | 1.017 | 0.542-1.908 |
| **IVH** | 0.159 | 0.327 | 0.63 | 1.172 | 0.618-2.223 |
| **Mother high school** | 0.054 | 0.263 | 0.84 | 1.056 | 0.630-1.769 |
| **Cerebral palsy** | -0.188 | 1.360 | 0.89 | 0.828 | 0.058-11.912 |
| **IQ** at age 5 years | -0.075 | 0.013 | <0.001 | 0.928 | 0.905-0.951 |
| **MABC** at age 5 years | -0.127 | 0.049 | 0.009 | 0.881 | 0.801-0.969 |
| **Clubsports** at age 5 years | -0.662 | 0.256 | 0.01 | 0.516 | 0.312-0.852 |
| **SDQ** at age 5 years | 0.133 | 0.022 | <0.001 | 1.143 | 1.093-1.194 |

Abbreviation:
BPD: Bronchopulmonary Dysplasia, IQ: Intelligence Quotient, IVH: Intraventricular Hemorrhage, MABC: Movement Assessment Battery for Children- II, SDQ: Strengths and Difficulties Questionnaire, S.E.: standard error, B: Regression coefficient, CI: Confidence interval

**Supplemental-Table S 26.** Clubsports

| | B | S.E. | p-Value | Odds Ratio | 95%-CI |
|---|---|---|---|---|---|
| **Delayed school entry** | -0.346 | 0.407 | 0.40 | 0.707 | 0.319-1.569 |
| **Gestational age** | -0.048 | 0.080 | 0.55 | 0.953 | 0.815-1.115 |
| **Birth weight** | 0.000 | 0.001 | 0.57 | 1.000 | 0.998-1.001 |
| **Male sex** | -0.389 | 0.224 | 0.08 | 0.678 | 0.437-1.051 |
| **BPD** | 0.640 | 0.329 | 0.05 | 1.896 | 0.996-3.611 |
| **IVH** | 0.101 | 0.313 | 0.75 | 1.106 | 0.598-2.043 |
| **Mother high school** | 0.521 | 0.219 | 0.02 | 1.684 | 1.096-2.587 |
| **Cerebral palsy** | -0.958 | 1.279 | 0.45 | 0.384 | 0.031-4.710 |
| **IQ** at age 5 years | 0.056 | 0.010 | <0.001 | 1.058 | 1.036-1.080 |
| **MABC** at age 5 years | 0.053 | 0.040 | 0.18 | 1.054 | 0.976-1.140 |
| **Clubsports** at age 5 years | 1.696 | 0.233 | <0.001 | 5.455 | 3.455-8.612 |
| **SDQ** at age 5 years | 0.016 | 0.021 | 0.46 | 1.016 | 0.975-1.059 |

Abbreviation:
BPD: Bronchopulmonary Dysplasia, IQ: Intelligence Quotient, IVH: Intraventricular Hemorrhage, MABC: Movement Assessment Battery for Children- II, SDQ: Strengths and Difficulties Questionnaire, S.E.: standard error, B: Regression coefficient, CI: Confidence interval

**Supplemental-Table S 27.** Participation (CASP, parental reported)

| | B | S.E. | p-Value | 95%-CI | |
|---|---|---|---|---|---|
| **Delayed school entry** | -1.037 | 0.999 | 0.30 | -2.997 | 0.923 |
| **Gestational age** | 0.022 | 0.164 | 0.90 | -0.300 | 0.343 |
| **Birth weight** | 0.001 | 0.001 | 0.55 | -0.002 | 0.003 |
| **Male sex** | 0.704 | 0.454 | 0.12 | -0.186 | 1.594 |
| **BPD** | 0.083 | 0.641 | 0.90 | -1.175 | 1.341 |
| **IVH** | 0.012 | 0.645 | 0.99 | -1.254 | 1.279 |
| **Mother high school** | -0.252 | 0.448 | 0.57 | -1.132 | 0.627 |
| **Cerebral palsy** | -7.359 | 3.799 | 0.05 | -14.815 | 0.097 |
| **IQ** at age 5 years | 0.081 | 0.021 | <0.001 | 0.040 | 0.122 |
| **MABC** at age 5 years | 0.349 | 0.080 | <0.001 | 0.193 | 0.506 |
| **Clubsports** at age 5 years | 1.008 | 0.448 | 0.03 | 0.129 | 1.886 |
| **SDQ** at age 5 years | -0.578 | 0.043 | <0.001 | -0.663 | -0.493 |

Abbreviation:
BPD: Bronchopulmonary Dysplasia, IQ: Intelligence Quotient, IVH: Intraventricular Hemorrhage, MABC: Movement Assessment Battery for Children- II, SDQ: Strengths and Difficulties Questionnaire, S.E.: standard error, B: Regression coefficient, CI: Confidence interval

**Supplemental-Table S 28.** Quality of life (KIDSCREEN-52, self-reported)

| | **B** | **S.E.** | **p-Value** | **95%-CI** | |
|---|---|---|---|---|---|
| **Delayed school entry** | 0.716 | 0.919 | 0.44 | -1.088 | 2.519 |
| **Gestational age** | -0.131 | 0.149 | 0.38 | -0.424 | 0.162 |
| **Birth weight** | 0.000 | 0.001 | 0.85 | -0.002 | 0.002 |
| **Male sex** | -0.593 | 0.418 | 0.16 | -1.413 | 0.227 |
| **BPD** | 0.232 | 0.593 | 0.70 | -0.931 | 1.395 |
| **IVH** | -0.865 | 0.584 | 0.14 | -2.012 | 0.281 |
| **Mother high school** | -0.006 | 0.413 | 0.99 | -0.817 | 0.805 |
| **Cerebral palsy** | 2.354 | 3.462 | 0.50 | -4.441 | 9.148 |
| **IQ** at age 5 years | 0.024 | 0.019 | 0.21 | -0.014 | 0.061 |
| **MABC** at age 5 years | 0.093 | 0.073 | 0.21 | -0.051 | 0.237 |
| **Clubsports** at age 5 years | 0.475 | 0.411 | 0.25 | -0.332 | 1.282 |
| **SDQ** at age 5 years | -0.338 | 0.040 | <0.001 | -0.417 | -0.260 |

Abbreviation:
BPD: Bronchopulmonary Dysplasia, IQ: Intelligence Quotient, IVH: Intraventricular Hemorrhage, MABC: Movement Assessment Battery for Children- II, SDQ: Strengths and Difficulties Questionnaire, S.E.: standard error, B: Regression coefficient, CI: Confidence interval

**Supplemental-Table S 29.** Bullying (KIDSCREEN-52, self-reported)

| | **B** | **S.E.** | **p-Value** | **95%-CI** | |
|---|---|---|---|---|---|
| **Delayed school entry** | 0.451 | 1.569 | 0.77 | -2.629 | 3.530 |
| **Gestational age** | -0.503 | 0.255 | 0.05 | -1.003 | -0.003 |
| **Birth weight** | 0.003 | 0.002 | 0.16 | -0.001 | 0.006 |
| **Male sex** | -0.378 | 0.714 | 0.60 | -1.778 | 1.023 |
| **BPD** | 0.815 | 1.012 | 0.42 | -1.171 | 2.802 |
| **IVH** | -2.357 | 0.998 | 0.02 | -4.315 | -0.400 |
| **Mother high school** | 0.377 | 0.706 | 0.59 | -1.009 | 1.762 |
| **Cerebral palsy** | 4.672 | 5.912 | 0.43 | -6.931 | 16.276 |
| **IQ** at age 5 years | 0.074 | 0.033 | 0.03 | 0.009 | 0.138 |
| **MABC** at age 5 years | 0.103 | 0.125 | 0.41 | -0.143 | 0.349 |
| **Clubsports** at age 5 years | -0.442 | 0.702 | 0.53 | -1.820 | 0.935 |
| **SDQ** at age 5 years | -0.451 | 0.068 | <0.001 | -0.585 | -0.316 |

Abbreviation:
BPD: Bronchopulmonary Dysplasia, IQ: Intelligence Quotient, IVH: Intraventricular Hemorrhage, MABC: Movement Assessment Battery for Children- II, SDQ: Strengths and Difficulties Questionnaire, S.E.: standard error, B: Regression coefficient, CI: Confidence interval

**Supplemental-Table S 30.** Attention (SDQ, self-reported)

| | **B** | **S.E.** | **p-Value** | **95%-CI** | |
|---|---|---|---|---|---|
| **Delayed school entry** | 0.077 | 0.167 | 0.64 | -0.251 | 0.406 |
| **Gestational age** | 0.016 | 0.027 | 0.56 | -0.038 | 0.070 |
| **Birth weight** | 0.000 | 0.000 | 0.14 | -0.001 | 0.000 |
| **Male sex** | -0.157 | 0.076 | 0.04 | -0.307 | -0.008 |
| **BPD** | -0.144 | 0.108 | 0.18 | -0.355 | 0.068 |
| **IVH** | 0.029 | 0.106 | 0.79 | -0.180 | 0.238 |
| **Mother high school** | -0.141 | 0.075 | 0.06 | -0.289 | 0.007 |
| **Cerebral palsy** | 0.120 | 0.630 | 0.85 | -1.117 | 1.357 |
| **IQ** at age 5 years | -0.012 | 0.003 | 0.001 | -0.018 | -0.005 |
| **MABC** at age 5 years | -0.024 | 0.013 | 0.08 | -0.050 | 0.002 |
| **Clubsports** at age 5 years | -0.016 | 0.075 | 0.83 | -0.164 | 0.131 |
| **SDQ** at age 5 years | 0.081 | 0.007 | <0.001 | 0.066 | 0.095 |

Abbreviation:
BPD: Bronchopulmonary Dysplasia, IQ: Intelligence Quotient, IVH: Intraventricular Hemorrhage, MABC: Movement Assessment Battery for Children- II, SDQ: Strengths and Difficulties Questionnaire, S.E.: standard error, B: Regression coefficient, CI: Confidence interval

**Supplemental-Table S 31.** Internalizing problems (SDQ, self-reported)

| | **B** | **S.E.** | **p-Value** | **95%-CI** | |
|---|---|---|---|---|---|
| **Delayed school entry** | 0.156 | 0.473 | 0.74 | -0.773 | 1.084 |
| **Gestational age** | 0.126 | 0.077 | 0.10 | -0.025 | 0.276 |
| **Birth weight** | -0.001 | 0.001 | 0.18 | -0.002 | 0.000 |
| **Male sex** | 0.847 | 0.215 | <0.001 | 0.425 | 1.269 |
| **BPD** | 0.245 | 0.305 | 0.42 | -0.354 | 0.844 |
| **IVH** | 0.504 | 0.301 | 0.09 | -0.086 | 1.094 |
| **Mother high school** | -0.012 | 0.213 | 0.95 | -0.430 | 0.405 |
| **Cerebral palsy** | -0.378 | 1.783 | 0.83 | -3.877 | 3.121 |
| **IQ** at age 5 years | -0.006 | 0.010 | 0.53 | -0.026 | 0.013 |
| **MABC** at age 5 years | -0.061 | 0.038 | 0.11 | -0.135 | 0.013 |
| **Clubsports** at age 5 years | -0.060 | 0.212 | 0.78 | -0.476 | 0.355 |
| **SDQ** at age 5 years | 0.241 | 0.021 | <0.001 | 0.200 | 0.281 |

Abbreviation:
BPD: Bronchopulmonary Dysplasia, IQ: Intelligence Quotient, IVH: Intraventricular Hemorrhage, MABC: Movement Assessment Battery for Children- II, SDQ: Strengths and Difficulties Questionnaire, S.E.: standard error, B: Regression coefficient, CI: Confidence interval

**Supplemental-Table S 32.** School (KOPKIJ, parental reported)

| | B | S.E. | p-Value | 95%-CI | |
|---|---|---|---|---|---|
| **Delayed school entry** | 0.120 | 0.072 | 0.10 | -0.021 | 0.261 |
| **Gestational age** | 0.017 | 0.012 | 0.16 | -0.007 | 0.041 |
| **Birth weight** | 0.000 | 0.000 | 0.45 | 0.000 | 0.000 |
| **Male sex** | 0.002 | 0.033 | 0.94 | -0.063 | 0.068 |
| **BPD** | -0.018 | 0.047 | 0.70 | -0.111 | 0.075 |
| **IVH** | -0.009 | 0.047 | 0.85 | -0.102 | 0.084 |
| **Mother high school** | -0.050 | 0.033 | 0.13 | -0.115 | 0.015 |
| **Cerebral palsy** | -0.459 | 0.289 | 0.11 | -1.026 | 0.108 |
| **IQ** at age 5 years | -0.011 | 0.002 | <0.001 | -0.014 | 0.008 |
| **MABC** at age 5 years | -0.017 | 0.006 | 0.004 | -0.029 | -0.006 |
| **Clubsports** at age 5 years | -0.055 | 0.033 | 0.10 | -0.120 | 0.010 |
| **SDQ** at age 5 years | 0.029 | 0.003 | <0.001 | 0.023 | 0.035 |

Abbreviation:
BPD: Bronchopulmonary Dysplasia, IQ: Intelligence Quotient, IVH: Intraventricular Hemorrhage, MABC: Movement Assessment Battery for Children- II, SDQ: Strengths and Difficulties Questionnaire, S.E.: standard error, B: Regression coefficient, CI: Confidence interval

**Supplemental-Table S 33.** Executive Function (BRIEF, parental reported)

| | B | S.E. | p-Value | 95%-CI | |
|---|---|---|---|---|---|
| **Delayed school entry** | 0.573 | 1.301 | 0.66 | -1.980 | 3.126 |
| **Gestational age** | 0.086 | 0.221 | 0.70 | -0.347 | 0.520 |
| **Birth weight** | -0.003 | 0.002 | 0.08 | -0.006 | 0.000 |
| **Male sex** | 1.038 | 0.610 | 0.09 | -0.158 | 2.234 |
| **BPD** | -2.374 | 0.862 | 0.006 | -4.066 | -0.682 |
| **IVH** | -0.712 | 0.860 | 0.41 | -2.399 | 0.974 |
| **Mother high school** | 0.390 | 0.604 | 0.52 | -0.796 | 1.576 |
| **Cerebral palsy** | 10.032 | 5.280 | 0.06 | -20.395 | 0.330 |
| **IQ** at age 5 years | -0.067 | 0.028 | 0.02 | -0.122 | -0.012 |
| **MABC** at age 5 years | -0.389 | 0.107 | <0.001 | -0.599 | -0.180 |
| **Clubsports** at age 5 years | -1.382 | 0.604 | 0.02 | -2.568 | -0.196 |
| **SDQ** at age 5 years | 1.067 | 0.058 | <0.001 | 0.953 | 1.182 |

Abbreviation:
BPD: Bronchopulmonary Dysplasia, IQ: Intelligence Quotient, IVH: Intraventricular Hemorrhage, MABC: Movement Assessment Battery for Children- II, SDQ: Strengths and Difficulties Questionnaire, S.E.: standard error, B: Regression coefficient, CI: Confidence interval

**Supplemental-Table S 34.** Speech (KOPKIJ, parental reported)

| | **B** | **S.E.** | **p-Value** | **95%-CI** | |
|---|---|---|---|---|---|
| **Delayed school entry** | 0.072 | 0.032 | 0.03 | 0.008 | 0.136 |
| **Gestational age** | 0.002 | 0.006 | 0.70 | -0.009 | 0.013 |
| **Birth weight** | 0.000 | 0.000 | 0.56 | 0.000 | 0.000 |
| **Male sex** | 0.003 | 0.015 | 0.85 | -0.027 | 0.033 |
| **BPD** | 0.005 | 0.022 | 0.82 | -0.037 | 0.047 |
| **IVH** | 0.004 | 0.021 | 0.85 | -0.038 | 0.046 |
| **Mother high school** | -0.027 | 0.015 | 0.08 | -0.056 | 0.003 |
| **Cerebral palsy** | -0.267 | 0.132 | 0.04 | -0.526 | -0.009 |
| **IQ** at age 5 years | -0.004 | 0.001 | <0.001 | -0.005 | -0.002 |
| **MABC** at age 5 years | -0.005 | 0.003 | 0.06 | -0.010 | 0.000 |
| **Clubsports** at age 5 years | -0.013 | 0.015 | 0.38 | -0.043 | 0.016 |
| **SDQ** at age 5 years | 0.011 | 0.001 | <0.001 | 0.008 | 0.014 |

Abbreviation:
BPD: Bronchopulmonary Dysplasia, IQ: Intelligence Quotient, IVH: Intraventricular Hemorrhage, MABC: Movement Assessment Battery for Children- II, SDQ: Strengths and Difficulties Questionnaire, S.E.: standard error, B: Regression coefficient, CI: Confidence interval

**Supplemental-Table S 35.** Memory (KOPKIJ, parental reported)

| | **B** | **S.E.** | **p-Value** | **95%-CI** | |
|---|---|---|---|---|---|
| **Delayed school entry** | 0.048 | 0.055 | 0.38 | -0.059 | 0.156 |
| **Gestational age** | 0.014 | 0.009 | 0.15 | -0.005 | 0.032 |
| **Birth weight** | 0.000 | 0.000 | 0.23 | 0.000 | 0.000 |
| **Male sex** | -0.025 | 0.026 | 0.33 | -0.076 | 0.025 |
| **BPD** | -0.035 | 0.036 | 0.34 | -0.107 | 0.036 |
| **IVH** | -0.022 | 0.036 | 0.54 | -0.093 | 0.049 |
| **Mother high school** | -0.037 | 0.026 | 0.15 | -0.087 | 0.013 |
| **Cerebral palsy** | -0.325 | 0.223 | 0.15 | -0.763 | 0.112 |
| **IQ** at age 5 years | -0.006 | 0.001 | <0.001 | -0.008 | -0.003 |
| **MABC** at age 5 years | -0.007 | 0.005 | 0.15 | -0.015 | 0.002 |
| **Clubsports** at age 5 years | -0.072 | 0.026 | 0.005 | -0.122 | -0.022 |
| **SDQ** at age 5 years | 0.034 | 0.002 | <0.001 | 0.029 | 0.038 |

Abbreviation:
BPD: Bronchopulmonary Dysplasia, IQ: Intelligence Quotient, IVH: Intraventricular Hemorrhage, MABC: Movement Assessment Battery for Children- II, SDQ: Strengths and Difficulties Questionnaire, S.E.: standard error, B: Regression coefficient, CI: Confidence interval

**Supplemental-Table S 36.** Visual-spatial (KOPKIJ, parental reported)

| | B | S.E. | p-Value | 95%-CI | |
|---|---|---|---|---|---|
| **Delayed school entry** | 0.118 | 0.064 | 0.06 | -0.007 | 0.244 |
| **Gestational age** | -0.014 | 0.011 | 0.20 | -0.035 | 0.007 |
| **Birth weight** | 0.000 | 0.000 | 0.61 | 0.000 | 0.000 |
| **Male sex** | -0.186 | 0.030 | <0.001 | -0.244 | -0.127 |
| **BPD** | 0.053 | 0.042 | 0.21 | -0.030 | 0.136 |
| **IVH** | 0.008 | 0.042 | 0.84 | -0.074 | 0.091 |
| **Mother high school** | 0.019 | 0.030 | 0.52 | -0.039 | 0.077 |
| **Cerebral palsy** | 0.907 | 0.259 | <0.001 | 0.399 | 1.415 |
| **IQ** at age 5 years | -0.007 | 0.001 | <0.001 | -0.010 | -0.004 |
| **MABC** at age 5 years | -0.017 | 0.005 | 0.002 | -0.027 | -0.006 |
| **Clubsports** at age 5 years | 0.031 | 0.030 | 0.30 | -0.027 | 0.089 |
| **SDQ** at age 5 years | 0.016 | 0.003 | <0.001 | 0.011 | 0.022 |

Abbreviation:
BPD: Bronchopulmonary Dysplasia, IQ: Intelligence Quotient, IVH: Intraventricular Hemorrhage, MABC: Movement Assessment Battery for Children- II, SDQ: Strengths and Difficulties Questionnaire, S.E.: standard error, B: Regression coefficient, CI: Confidence interval

**Supplemental-Table S 37.** Parents (KISCREEN-52, self reported)

| | B | S.E. | p-Value | 95%-CI | |
|---|---|---|---|---|---|
| **Delayed school entry** | 0.002 | 1.177 | 1.00 | -2.309 | 2.313 |
| **Gestational age** | -0.175 | 0.193 | 0.37 | -0.553 | 0.204 |
| **Birth weight** | 0.000 | 0.001 | 0.73 | -0.002 | 0.003 |
| **Male sex** | -0.031 | 0.540 | 0.95 | -1.091 | 1.029 |
| **BPD** | 0.293 | 0.765 | 0.70 | -1.207 | 1.794 |
| **IVH** | -0.226 | 0.755 | 0.77 | -1.707 | 1.255 |
| **Mother high school** | -0.405 | 0.534 | 0.45 | -1.453 | 0.643 |
| **Cerebral palsy** | 1.469 | 4.475 | 0.74 | -7.314 | 10.252 |
| **IQ** at age 5 years | 0.015 | 0.025 | 0.55 | -0.034 | 0.063 |
| **MABC** at age 5 years | 0.051 | 0.095 | 0.59 | -0.135 | 0.237 |
| **Clubsports** at age 5 years | 0.993 | 0.531 | 0.06 | -0.049 | 2.035 |
| **SDQ** at age 5 years | -0.251 | 0.052 | <0.001 | -0.353 | -0.149 |

Abbreviation:
BPD: Bronchopulmonary Dysplasia, IQ: Intelligence Quotient, IVH: Intraventricular Hemorrhage, MABC: Movement Assessment Battery for Children- II, SDQ: Strengths and Difficulties Questionnaire, S.E.: standard error, B: Regression coefficient, CI: Confidence interval

**Supplemental-Table S 38.** Peers (KIDSCREEN-52, self-reported)

| | **B** | **S.E.** | **p-Value** | **95%-CI** | |
|---|---|---|---|---|---|
| **Delayed school entry** | 0.213 | 1.483 | 0.89 | -2.698 | 3.125 |
| **Gestational age** | -0.233 | 0.241 | 0.33 | -0.706 | 0.239 |
| **Birth weight** | 0.000 | 0.002 | 0.86 | -0.003 | 0.004 |
| **Male sex** | -0.180 | 0.675 | 0.79 | -1.505 | 1.144 |
| **BPD** | 0.459 | 0.957 | 0.63 | -1.419 | 2.337 |
| **IVH** | -0.620 | 0.943 | 0.51 | -2.471 | 1.231 |
| **Mother high school** | -0.196 | 0.667 | 0.77 | -1.505 | 1.114 |
| **Cerebral palsy** | -1.394 | 5.589 | 0.80 | -12.365 | 9.576 |
| **IQ** at age 5 years | 0.039 | 0.031 | 0.21 | -0.022 | 0.099 |
| **MABC** at age 5 years | 0.228 | 0.118 | 0.05 | -0.004 | 0.461 |
| **Clubsports** at age 5 years | 1.085 | 0.664 | 0.10 | -0.218 | 2.387 |
| **SDQ** at age 5 years | -0.431 | 0.065 | <0.001 | -0.558 | -0.304 |

Abbreviation:
BPD: Bronchopulmonary Dysplasia, IQ: Intelligence Quotient, IVH: Intraventricular Hemorrhage, MABC: Movement Assessment Battery for Children- II, SDQ: Strengths and Difficulties Questionnaire, S.E.: standard error, B: Regression coefficient, CI: Confidence interval

**Supplemental-Table S 39.** Autonomy (KIDSCREEN-52, parental reported)

| | **B** | **S.E.** | **p-Value** | **95%-CI** | |
|---|---|---|---|---|---|
| **Delayed school entry** | 2.764 | 1.175 | 0.02 | 0.459 | 5.069 |
| **Gestational age** | -0.002 | 0.193 | 0.99 | -0.380 | 0.376 |
| **Birth weight** | -0.001 | 0.001 | 0.39 | -0.004 | 0.002 |
| **Male sex** | -0.827 | 0.533 | 0.12 | -1.873 | 0.219 |
| **BPD** | -0.084 | 0.753 | 0.91 | -1.563 | 1.395 |
| **IVH** | -0.734 | 0.759 | 0.33 | -2.223 | 0.755 |
| **Mother high school** | -0.659 | 0.527 | 0.21 | -1.693 | 0.374 |
| **Cerebral palsy** | -8.420 | 4.469 | 0.06 | -17.191 | 0.351 |
| **IQ** at age 5 years | -0.038 | 0.025 | 0.12 | -0.087 | 0.010 |
| **MABC** at age 5 years | 0.223 | 0.094 | 0.02 | 0.039 | 0.407 |
| **Clubsports** at age 5 years | -0.582 | 0.526 | 0.27 | -1.614 | 0.450 |
| **SDQ** at age 5 years | -0.324 | 0.051 | <0.001 | -0.424 | -0.224 |

Abbreviation:
BPD: Bronchopulmonary Dysplasia, IQ: Intelligence Quotient, IVH: Intraventricular Hemorrhage, MABC: Movement Assessment Battery for Children- II, SDQ: Strengths and Difficulties Questionnaire, S.E.: standard error, B: Regression coefficient, CI: Confidence interval

**Appendix 6. Total regression analysis: Timely transition to secondary school (corrected age).**

**Supplemental-Table S 40.** Primary school (Early school entry vs. age-appropriate school entry)

| | **B** | **S.E.** | **p-Value** | **Odds Ratio** | **95%-CI** |
|---|---|---|---|---|---|
| **age-appropriate school entry** | -1.104 | 0.264 | <0.001 | 0.332 | 0.197-0.557 |
| **Gestational age** | -0.049 | 0.094 | 0.60 | 0.952 | 0.792-1.145 |
| **Birth weight** | 0.000 | 0.001 | 0.97 | 1.000 | 0.999-1.001 |
| **Male sex** | 0.044 | 0.263 | 0.87 | 1.045 | 0.624-1.752 |
| **BPD** | -0.372 | 0.409 | 0.36 | 0.689 | 0.309-1.537 |
| **IVH** | 0.296 | 0.372 | 0.43 | 1.344 | 0.649-2.786 |
| **Mother high school** | -0.403 | 0.262 | 0.12 | 0.668 | 0.400-1.116 |
| **Cerebral palsy** | n too small for calculation | | | | |
| **IQ** at age 5 years | -0.046 | 0.012 | <0.001 | 0.955 | 0.933-0.977 |
| **MABC** at age 5 years | 0.026 | 0.047 | 0.58 | 1.026 | 0.935-1.126 |
| **Clubsports** at age 5 years | -0.023 | 0.267 | 0.93 | 0.977 | 0.579-1.648 |
| **SDQ** at age 5 years | 0.018 | 0.025 | 0.02 | 1.018 | 0.970-1.069 |

Abbreviation:
BPD: Bronchopulmonary Dysplasia, IQ: Intelligence Quotient, IVH: Intraventricular Hemorrhage, MABC: Movement Assessment Battery for Children- II, SDQ: Strengths and Difficulties Questionnaire, S.E.: standard error, B: Regression coefficient, CI: Confidence interval

**Supplemental-Table S 41.** Secondary school (Early school entry vs. age-appropriate school entry)

| | B | S.E. | p-Value | Odds Ratio | 95%-CI |
|---|---|---|---|---|---|
| **age-appropriate school entry** | 1.023 | 0.251 | <0.001 | 2.781 | 1.700-4.550 |
| **Gestational age** | 0.022 | 0.091 | 0.81 | 1.022 | 0.856-1.221 |
| **Birth weight** | 0.000 | 0.001 | 0.81 | 1.000 | 0.999-1.002 |
| **Male sex** | -0.110 | 0.254 | 0.67 | 0.896 | 0.545-1.474 |
| **BPD** | 0.273 | 0.387 | 0.48 | 1.314 | 0.615-2.806 |
| **IVH** | -0.376 | 0.368 | 0.31 | 0.687 | 0.334-1.411 |
| **Mother high school** | 0.252 | 0.251 | 0.31 | 1.287 | 0.787-2.103 |
| **Cerebral palsy** | -0.503 | 1.564 | 0.75 | 0.605 | 0.028-12.963 |
| **IQ** at age 5 years | 0.063 | 0.012 | <0.001 | 1.065 | 1.041-1.090 |
| **MABC** at age 5 years | -0.025 | 0.046 | 0.60 | 0.976 | 0.891-1.068 |
| **Clubsports** at age 5 years | 0.059 | 0.257 | 0.82 | 1.061 | 0.641-1.755 |
| **SDQ** at age 5 years | -0.029 | 0.024 | 0.23 | 0.972 | 0.927-1.019 |

Abbreviation:
BPD: Bronchopulmonary Dysplasia, IQ: Intelligence Quotient, IVH: Intraventricular Hemorrhage, MABC: Movement Assessment Battery for Children- II, SDQ: Strengths and Difficulties Questionnaire, S.E.: standard error, B: Regression coefficient, CI: Confidence interval

**Supplemental-Table S 5.** Secondary school (age-appropriate school entry vs. delayed school entry)

| | B | S.E. | p-Value | Odds Ratio | 95%-CI |
|---|---|---|---|---|---|
| **delayed school entry** | 0.708 | 0.979 | 0.47 | 2.031 | 0.298-13.820 |
| **Gestational age** | 0.073 | 0.088 | 0.40 | 1.076 | 0.906-1.278 |
| **Birth weight** | 0.000 | 0.001 | 0.98 | 1.00 | 0.999-1.001 |
| **Male sex** | -0.184 | 0.246 | 0.46 | 0.832 | 0.513-1.349 |
| **BPD** | 0.246 | 0.368 | 0.50 | 1.279 | 0.622-2.630 |
| **IVH** | -0.419 | 0.348 | 0.23 | 0.658 | 0.333-1.300 |
| **Mother high school** | 0.279 | 0.242 | 0.25 | 1.322 | 0.832-2.124 |
| **Cerebral palsy** | -0.155 | 1.511 | 0.92 | 0.857 | 0.044-16.542 |
| **IQ** at age 5 years | 0.056 | 0.011 | <0.001 | 1.058 | 1.035-1.081 |
| **MABC** at age 5 years | -0.021 | 0.045 | 0.64 | 0.979 | 0.896-1.070 |
| **Clubsports** at age 5 years | 0.019 | 0.247 | 0.94 | 1.019 | 0.628-1.655 |
| **SDQ** at age 5 years | -0.039 | 0.024 | 0.10 | 0.962 | 0.919-1.007 |

Abbreviation:
BPD: Bronchopulmonary Dysplasia, IQ: Intelligence Quotient, IVH: Intraventricular Hemorrhage, MABC: Movement Assessment Battery for Children- II, SDQ: Strengths and Difficulties Questionnaire, S.E.: standard error, B: Regression coefficient, CI: Confidence interval

**Appendix 7. Total regression analysis: Timely transition to secondary school (chronological age).**

**Supplemental-Table S 43.** Primary school (Early school entry vs. age-appropriate school entry)

| | **B** | **S.E.** | **p-Value** | **Odds Ratio** | **95%-CI** |
|---|---|---|---|---|---|
| **age-appropriate school entry** | -1.073 | 0.257 | <0.001 | 0.342 | 0.207-0.566 |
| **Gestational age** | -0.075 | 0.095 | 0.43 | 0.928 | 0.770-1.118 |
| **Birth weight** | 0.000 | 0.001 | 0.92 | 1.000 | 0.998-1.001 |
| **Male sex** | 0.057 | 0.264 | 0.83 | 1.058 | 0.631-1.775 |
| **BPD** | -0.455 | 0.412 | 0.27 | 0.634 | 0.283-1.422 |
| **IVH** | 0.440 | 0.371 | 0.24 | 1.553 | 0.751-3.213 |
| **Mother high school** | -0.308 | 0.264 | 0.24 | 0.735 | 0.438-1.234 |
| **Cerebral palsy** | n too small for calculation | | | | |
| **IQ** at age 5 years | -0.042 | 0.012 | <0.001 | 0.959 | 0.937-0.981 |
| **MABC** at age 5 years | 0.007 | 0.048 | 0.88 | 1.007 | 0.918-1.106 |
| **Clubsports** at age 5 years | -0.003 | 0.268 | 0.99 | 0.997 | 0.590-1.685 |
| **SDQ** at age 5 years | 0.012 | 0.026 | 0.65 | 1.012 | 0.962-1.065 |

Abbreviation:

BPD: Bronchopulmonary Dysplasia. IQ: Intelligence Quotient. IVH: Intraventricular Hemorrhage. MABC: Movement Assessment Battery for Children- II. SDQ: Strengths and Difficulties Questionnaire. S.E.: standard error. B: Regression coefficient. CI: Confidence interval

**Supplemental-Table S 44.** Secondary school (Early school entry vs. age-appropriate school entry)

| | B | S.E. | p-Value | Odds Ratio | 95%-CI |
|---|---|---|---|---|---|
| **age-appropriate school entry** | 0.935 | 0.252 | <0.001 | 2.548 | 1.554-4.179 |
| **Gestational age** | 0.053 | 0.091 | 0.56 | 1.055 | 0.883-1.260 |
| **Birth weight** | 0.000 | 0.001 | 0.85 | 1.000 | 0.999-1.002 |
| **Male sex** | -0.124 | 0.254 | 0.63 | 0.884 | 0.537-1.454 |
| **BPD** | 0.367 | 0.389 | 0.35 | 1.443 | 0.673-3.094 |
| **IVH** | -0.507 | 0.366 | 0.17 | 0.602 | 0.294-1.233 |
| **Mother high school** | 0.152 | 0.253 | 0.55 | 1.164 | 0.709-1.911 |
| **Cerebral palsy** | -0.464 | 1.524 | 0.76 | 0.629 | 0.032-12.481 |
| **IQ** at age 5 years | 0.059 | 0.012 | <0.001 | 01.061 | 1.037-1.085 |
| **MABC** at age 5 years | -0.008 | 0.046 | 0.87 | 0.992 | 0.906-1.087 |
| **Clubsports** at age 5 years | 0.048 | 0.257 | 0.85 | 1.050 | 0.634-1.739 |
| **SDQ** at age 5 years | -0.026 | 0.025 | 0.29 | 0.974 | 0.927-1.023 |

Abbreviation:
BPD: Bronchopulmonary Dysplasia. IQ: Intelligence Quotient. IVH: Intraventricular Hemorrhage. MABC: Movement Assessment Battery for Children- II. SDQ: Strengths and Difficulties Questionnaire. S.E.: standard error. B: Regression coefficient. CI: Confidence interval

**Supplemental-Table S 45.** Secondary school (age-appropriate school entry vs. delayed school entry)

| | B | S.E. | p-Value | Odds Ratio | 95%-CI |
|---|---|---|---|---|---|
| **delayed school entry** | 1.794 | 0.837 | 0.03 | 6.011 | 1.166-31.000 |
| **Gestational age** | 0.091 | 0.088 | 0.30 | 1.095 | 0.921-1.303 |
| **Birth weight** | 0.000 | 0.001 | 0.88 | 1.000 | 0.999-1.001 |
| **Male sex** | -0.159 | 0.247 | 0.52 | 0.853 | 0.525-1.385 |
| **BPD** | 0.243 | 0.374 | 0.52 | 1.276 | 0.613-2.654 |
| **IVH** | -0.470 | 0.352 | 0.18 | 0.625 | 0.313-1.246 |
| **Mother high school** | 0.198 | 0.245 | 0.42 | 1.219 | 0.754-1.973 |
| **Cerebral palsy** | 0.125 | 1.514 | 0.93 | 1.133 | 0.058-22.043 |
| **IQ** at age 5 years | 0.056 | 0.011 | <0.001 | 1.058 | 1.035-1.081 |
| **MABC** at age 5 years | -0.016 | 0.046 | 0.73 | 0.984 | 0.900-1.076 |
| **Clubsports** at age 5 years | 0.072 | 0.251 | 0.77 | 1.075 | 0.658-1.757 |
| **SDQ** at age 5 years | -0.043 | 0.024 | 0.07 | 0.958 | 0.914-1.003 |

Abbreviation:
BPD: Bronchopulmonary Dysplasia. IQ: Intelligence Quotient. IVH: Intraventricular Hemorrhage. MABC: Movement Assessment Battery for Children- II. SDQ: Strengths and Difficulties Questionnaire. S.E.: standard error. B: Regression coefficient. CI: Confidence interval
